# Psychometric Properties of the AASPIRE Autistic Burnout Measure – Revised

**DOI:** 10.64898/2026.08.19.26360826

**Authors:** Christina Nicolaidis, Liu-Qin Yang, Mathew C. Uretsky, Dora Raymaker, Mary Baker-Ericzén, Vivian Darlene Grillo, Steven K. Kapp, Rachel Kripke-Ludwig, Joelle Maslak, Ian Moura, Mirah Scharer, Anna Furra Wallington

## Abstract

**Background:** Autistic Chronic Energy Depletion Syndrome, commonly referred to as Autistic Burnout, is a debilitating condition characterized by exhaustion, loss of function, and reduced tolerance to stimuli. While several instruments attempt to measure it, validation studies have only used cross-sectional designs and/or convenience samples with low support needs, limiting understanding of their performance across heterogeneous, autistic populations and over time.

**Methods:** Using a community-based participatory research (CBPR) approach, we revised the 27-item AASPIRE Autistic Burnout Measure (AABM) into a 14-item AASPIRE Autistic Burnout Measure-Revised (AABM-R) and tested it in a longitudinal study of 835 autistic adults recruited from healthcare systems, disability services, and the community. Participants completed surveys directly (with or without support) or via a caregiver. We assessed structural validity and measurement invariance using exploratory and confirmatory factor analysis, tested construct validity through a priori hypothesis testing, examined discriminant validity from depression using longitudinal factor analysis, and assessed criterion validity using ROC analysis.

**Results:** The AABM-R demonstrated a clear single-factor structure among direct reporters, with and without support, and measurement invariance across these groups; findings were less conclusive for the smaller caregiver-report subsample. Autistic burnout correlated as hypothesized with stressors (e.g., discrimination, masking, adverse childhood experiences), supports (e.g., social support, receiving needed help with daily living activities), and broader outcomes (e.g., quality of life, depression, anxiety). Longitudinal modeling supported autistic burnout as empirically distinct from, though related to, depression. ROC analysis (AUC = 0.89) supported cut-offs distinguishing probable (33-56, LR 7.38), unsure, and unlikely (0-22, LR 0.15) burnout.

**Conclusions:** The AABM-R is a brief, accessible, psychometrically sound measure of autistic burnout suitable for heterogeneous autistic populations, with preliminary clinical cut-offs to guide screening. Further research is needed on the caregiver-report version and on longitudinal predictors and outcomes of burnout.

**Community Brief:** *Why is this an important issue?:* Autistic burnout makes people feel exhausted, and it makes it harder for them to function or deal with sensory sensitivities. It can be a very serious problem. The autistic community has recognized autistic burnout for a long time, but the science is only starting to catch up. We need good ways to measure burnout to better understand and treat it, but researchers haven’t yet fully tested survey measures in diverse groups of autistic adults.

*What was the purpose of this study?:* To test a shortened, more accessible autistic burnout questionnaire called the AASPIRE Autistic Burnout Measure-Revised (AABM-R).

*What did the researchers do?:* Our team of autistic community members and researchers worked together as equal partners throughout the project. We shortened an earlier version of the questionnaire from 27 to 14 questions and tried to make it easier to understand. We tested this new questionnaire with 835 autistic adults. People in the sample had a wide range of abilities and experiences. Some people used the “direct report” version to answer questions themselves, with or without support. When that wasn’t possible, caregivers used the “caregiver report” version to answer questions for them. People took the same survey three times over about a year. We used a lot of statistics to see if the burnout scale works well.

*What were the results and conclusions of the study?:* 1) The direct report version of the AABM-R had good “structural validity.” That means it holds together well and it measures one single idea. The data was less clear for the caregiver report version. 2) For people using the direct report version, it worked the same way whether or not they needed help to take part in the study. (That’s called “measurement invariance.”) 3) The scale had good “construct validity.” That means that burnout scores were linked to stressors, supports, health, and quality of life in the ways we predicted. 4) The burnout scale measures something that is distinctly different from depression. 5) Scores of 0 to 22 on the AABM_R14 mean that burnout is less likely and scores of 33 to 56 mean that it is more likely. Scores in between don’t tell us much.

*What is new or controversial about these findings?:* The findings give us more reason to believe that the direct report version of the AABM-R works well and they help us interpret scores. Our study included autistic people with a wider range of strengths and challenges than other studies.

*What are potential weaknesses in the study?:* Not enough caregivers took part to see how well the caregiver-report version of the AABM-R works. We only measured masking with only one question, not a full questionnaire.

*How will these findings help autistic adults now or in the future?:* The AABM-R gives people a short way to measure autistic burnout. That can help guide conversations with clinicians about burnout. In the future, it may also help researchers understand what causes burnout and whether services and supports actually help.

## Introduction

Even though the autistic community has long recognized autistic burnout and its impacts on health and wellbeing, the concept only entered the clinical and academic literature relatively recently.^1,2^ Reliable and valid measurements of autistic burnout are essential not only for estimating prevalence and identifying individuals experiencing burnout, but also for advancing etiological research, evaluating interventions, and tracking recovery. Researchers have started to create and validate survey instruments to measure autistic burnout,^3–6^ but additional work is needed to refine such instruments and test their ability to accurately and efficiently screen for autistic burnout in autistic people with heterogeneous characteristics and to track changes in the severity of burnout over time.

### Conceptualizations of Autistic Burnout

Multiple research groups have independently arrived at similar descriptions of autistic burnout, using a variety of qualitative, survey, and consensus-forming methodologies. The Academic Autism Spectrum Partnership in Research and Education (AASPIRE) conducted a qualitative study, using a community based participatory research (CBPR) approach, to first describe autistic burnout. They characterized autistic burnout as “pervasive, long-term (typically 3+ months) exhaustion, loss of function, and reduced tolerance to stimulus.”^1^ Similarly, Higgins and colleagues used a grounded Delphi process to create a clinical definition of autistic burnout, describing it as a highly debilitating condition with significant mental and physical exhaustion, interpersonal withdrawal, and one or more of the following; 1) significant reduction in social, occupational, educational, academic, behavioral, or other important areas of functioning; 2) confusion, difficulties with executive function, and/or dissociative states; or 3) increased intensity of autistic traits and/or reduced capacity to camouflage/mask.^7^ There is general concordance between conceptualizations, other than the duration criteria for the condition, with the Higgins definition including brief or intermittent episodes,^8^ while the AASPIRE group defines burnout as a chronic condition, using an arbitrary cut-off of three months to match the clinical definition of other chronic conditions such as chronic pain, long COVID, or professional burnout.^1^

Research groups have also independently found very similar attributions for autistic burnout, all suggesting that it may represent an autism-specific manifestation of prolonged demand-resource imbalance.^1,7,9–11^ A scoping review recognized chronic masking, discrimination, adverse childhood experiences, significant life stressors, and structural barriers to healthcare and disability supports as contributing factors to autistic burnout.^2^ The small but growing body of empirical work suggests that autistic burnout is distinct from, though often co-occurring with, occupational burnout, depression, anxiety, or physical or mental health conditions.^1,4,6,9^

Emerging research suggests that autistic burnout may be associated with substantial impairment and reduced well-being. Qualitative and Delphi studies consistently show that autistic adults describe burnout as contributing to severe reductions in health and social functioning and quality of life.^1,2,7,10^ Moreover, cross-sectional studies have found associations between burnout and withdrawal from education and employment,^12^ increased depression,^9^ diminished flourishing, and reduced quality of life.^6^ However, more robust understandings of the predictors and consequences of autistic burnout require longitudinal analyses with sound measurement tools.

### Measurement of Autistic Burnout

In 2019, the AASPIRE Autistic Burnout team, led by autistic researcher Dr. Dora Raymaker, used a CBPR approach with clinicians and autistic community partners to create the AASPIRE Autistic Burnout Measure (AABM). The original AABM consisted of 27 items based on the team’s previous qualitative research.^1^ They began evaluating the psychometric properties of the AABM with promising preliminary findings. However, they halted data collection with the onset of the COVID-19 pandemic given concerns that pandemic-related factors would affect data collection and analysis. Based on the relatively small sample size (N=80), the high proportion of participants who had experienced burnout (96%), and expert feedback about potential accessibility issues raised in a separate project, they decided not to publish those preliminary results, but to continue to refine the measure (preprint of initial results available as Raymaker et al., 2026^3^).

Several groups used and tested the pre-publication version of the AABM, with findings supporting its internal consistency reliability, construct validity, and structural coherence.^4–6^ Dutch^13^ and Polish^14^ translations also showed promising psychometric properties. Another tool, the Autistic Burnout Severity Items (ABSI),^4^ also showed promising properties, but its structure only allows it to be used with participants who already believe they have or have had autistic burnout.

Despite promising initial testing, many questions remain. First, almost all these studies have primarily included convenience samples of autistic adults recruited via the internet, with most participants having been diagnosed in adulthood and/or having high educational attainment and relatively low support needs.^3–6,13^ Similarly, most prior studies have either limited their samples to people with prior experience of autistic burnout^4^ or have included samples with high proportions of people who had experienced autistic burnout, potentially due to the use of convenience samples recruited with materials mentioning burnout or camouflaging.^3,6^

Second, cross-sectional designs predominate, thus restricting understanding of longitudinal trajectories or the assessment of instruments’ test-retest reliability and responsiveness to change. Third, although construct validity has been partially established through correlations with depression, health, and functioning, modeling of both precursors and outcomes within a unified analytic framework remains limited.^2^ Finally, studies have not formally tested measurement invariance across different participation modalities (e.g., independent self-report versus supported self-report versus caregiver-report). Establishing such invariance ensures that burnout scores are interpretable and comparable across autistic adults with varying communication profiles and support needs.

Addressing these gaps is critical. Without psychometrically sound, accessible, and inclusive tools tested in heterogeneous, longitudinal samples, research on autistic burnout risks underrepresenting or mischaracterizing important subgroups of autistic individuals and limits its applicability for screening, service planning, and policy development.

### The Current Study

The present study is part of the larger AASPIRE Outcomes Project, which aimed to develop and test the AASPIRE Measurement Toolkit (www.aaspire.org/measurement), a set of accessible, co-produced, self-reported outcome measures that can be used to assess the effectiveness of services for autistic adults.

In the first phase of the project, we used a CBPR-nested Delphi process with 54 people, including external experts with lived and professional experience and our own community-academic team. All internal and external experts reached consensus on which outcomes were most important to measure.^15^ The participants also provided detailed feedback on how existing instruments, including the original AABM, would need to be changed to be used with autistic adults with heterogeneous demographic and disability characteristics. In phase two, we used our CBPR instrument adaptation process^16^ to collaboratively create or adapt 19 outcome measures focused on these high-priority outcomes, as well as 12 modules assessing detailed participant characteristics. Each instrument has two versions, one for use by autistic adults who can take part directly, either with or without support; and another for caregivers taking part on behalf of an autistic participant who cannot take part directly, even with accommodations and supports. All instruments are freely available at www.aaspire.org/measurment.

In phase three, we conducted a longitudinal study with 870 autistic adults residing in the United States, recruited from two healthcare systems, two disability services systems, and the broader autistic community.^17^ Participants could take part directly independently (DR sample) or with support (DR-S sample) or via a caregiver (CR sample). They took a survey at 3 time points over a 12-18-month period. A random 15% sample took an additional survey two weeks after the second time point to assess test-retest reliability.

Autistic burnout was one of the outcomes identified in the CBPR-nested Delphi process as being a high priority.^15^ Our community-academic team collaboratively revised the 27-item AABM to create the new 14-item AASPIRE Autistic Burnout Measure-Revised (AABM-R) and included the instrument in the larger longitudinal study. We present initial psychometric properties of all 19 outcome measures, including the AABM-R, elsewhere.^18^ Those analyses provide strong evidence for the AABM-R’s accessibility, content validity, internal consistency reliability, 2-week test-retest reliability, convergent validity with other health and social outcomes, and 6-month responsiveness to change, though results are limited for the caregiver report version due to the small N using that version.^18^ This paper provides 1) additional details about the AABM-R’s development, 2) more in-depth psychometric analyses to further understand and support construct validity, including assessment of structural validity, measurement invariance, additional a-priori hypothesis testing, and discriminant validity with depression; and 3) preliminary assessment of criterion validity and recommendations for scoring interpretation.

## Methods

### Community-Academic Partnership and Participatory Approach

We used a CBPR approach throughout all phases of the project. Details of our long-standing partnership,^19^ collaboration guidelines,^20^ and instrument adaptation processes^16^ are presented elsewhere, as are educational tools and examples (www.aaspire.org/inclusion-toolkit).

The academic side of the team included multiple autistic and non-autistic academic investigators, consultants, students, and staff, while the community side of the team included ten non-academic autistic community partners, a representative of a family-focused community-organization, and a representative of a governmental developmental disability services agency. An autistic Community Project Lead helped bridge the two sides. Multiple team members held additional roles as clinicians, service providers, and family members of autistic adults. Greater details about the Outcome Project team members are available elsewhere.^15^

The majority of academic and community team members had experienced autistic burnout themselves and/or had supported people with autistic burnout. We made all important decisions using a consensus process^19^ and strove to treat academic and community team members as equal partners throughout the project. All team members had the opportunity to co-author this paper.

### Instrument adaptation and cognitive interviewing

Overall, participants in the CBPR-nested Delphi process felt that the original AABM measured the intended construct well and recommended its use in the project. However, some participants raised concerns about its accessibility, especially for autistic adults with intellectual disability. Others recommended shortening it to make it more feasible for use in clinical, services, or other real-world settings.

A smaller Autistic Burnout workgroup, consisting of both autistic and non-autistic team members, used this feedback to create a first draft of the revised measure and then brought it to the full team for review, making additional changes as needed. Adaptations from the original AABM included 1) shortening the instrument to 14 items, 2) adding text to clarify the timeframe, 3) making minor wording changes to increase accessibility, 4) adding detailed descriptions and vignettes to more clearly define concepts, and 5) adding items about the onset of symptoms and participant attributions.

Once the full-team reached consensus on the revised draft, we included it in a cognitive interview study with 37 autistic adults and supporters. Participants who reviewed the AABM-R (6 autistic participants and 2 caregivers) were able to paraphrase items well and felt they covered the key aspects of autistic burnout, but some of the first participants forgot that the question prompt asked them to focus on symptoms that had been occurring for at least 3 months. As such, we decided to repeat the prompt in every item, which helped the remaining participants. The full academic-community team approved the final version (available as **Supplement A**).

### Longitudinal Survey Participants

All participants needed to be at least 18 years of age, live in the United States, identify as autistic, and either have the ability to take part in a survey in English themselves, with or without support, or have a caregiver who was able to take part on their behalf. Additionally, participants in the Healthcare subcohort had to receive healthcare services in one of two partnering healthcare systems in Portland, Oregon or Nashville, Tennessee and have a diagnostic code related to autism in their medical records. Participants in the Disability Services subcohort had to receive disability services for autism from governmental disability agencies in Multnomah County, Oregon or San Diego County, California. A detailed description of the overall study methods is available elsewhere.^17^

This analysis is limited to the 835 participants who completed the autistic burnout measure at baseline. Over 80% of participants who took part in the baseline survey participated in the follow-up surveys. Participants were divided relatively equally between the disability services, healthcare, and community subcohorts (31%, 36%, and 33% respectively). Almost all took part directly; 585 (70%) could do so without support and 180 (22%) required support; 70 (8%) took part via a caregiver. Age ranged from 18-77 years (mean 31; STD 11.6) and ∼30% identified as Black, Indigenous, or other People of Color (BIPOC). Over half the sample had challenges with spoken language and a majority (71%) needed at least some support with basic or instrumental activities of daily living. Approximately half were diagnosed with autism in childhood, and a majority had at least one co-occurring mental health condition. The sample included substantial heterogeneity in gender, sex assigned at birth, living arrangement, educational attainment, employment, and health status. Approximately half the participants had not heard of or did not know much about autistic burnout prior to taking the survey. Approximately half felt they likely had had autistic burnout at some point in their lives and a third felt they were in an episode of autistic burnout currently (**Supplement B**).

### Procedures

The Institutional Review Board (IRB) at Portland State University (PSU) approved this study. The IRBs at all partnering sites waved authorization to PSU. After a brief screening survey, participants or their legally authorized representatives offered informed consent using our accessible consent and authorization procedures^21^ and data sharing protocols.^22^

Participants could take the surveys online, via Zoom video conference, over the telephone, or where feasible, in person. They could receive support from their own supporters or a research assistant, as needed. We used the Research Electronic Data Capture (REDCap) system^23,24^ hosted at Oregon Health and Science University for data collection and management. The system included multiple accessibility features, such as hover text to define potentially confusing terms or provide examples. Detailed descriptions of our recruitment, screening, data collection, scammer prevention, and data integrity methods are available elsewhere.^17^

### Data Collection

The AABM-R includes 14 items about autistic burnout. Each uses a 0-4 Likert scale (0 = Strongly Disagree to 4 = Strongly Agree). The total score ranges from 0-56, with higher scores indicating greater severity of autistic burnout. For scale-level analyses, we computed mean AABM-R score by dividing the total score by the number of items answered, resulting in a range of 0-4. We used summed scores for ROC analyses and scoring interpretation. We used item-level data for factor-analytic and discriminant-validity analyses. The instrument also includes a detailed preface and additional items about symptom onset, participants’ attributions, prior familiarity with autistic burnout, and participants self-assessment of whether they have autistic burnout (**Supplement A**).

We collected information using the other 18 outcome instruments developed or adapted for the AASPIRE Outcome Measurement Toolkit, including measures of health and global life outcomes (quality of life; overall physical, mental, and social wellbeing; flourishing; health interference; depression; and anxiety); social outcomes (self-determination, choices and decisions, community participation, and two measures of employment satisfaction); supports (adequacy of information and supports, satisfaction with disability services, social support, and barriers to communication); and healthcare services (unmet healthcare needs, barriers to healthcare, patient-provider communication, and healthcare self-efficacy). All instruments are available at www.aaspire.org/measurement. Previously published results provide evidence to suggest that all outcome measures were accessible and demonstrated strong initial psychometric properties.^18^

We also used 12 modules to collect detailed information on participant characteristics and potential predictors. These modules included an item on major life events over the past 6 months, using a checklist for events such as major illness or injury or changes in employment, relationship status, or residence (see **Supplement B**). We dichotomized responses to those who had any major life events vs no major life events in the last 6 months. There were two items on the need for help with basic activities of daily living (ADLs) and instrumental activities of daily living (IADLs). For those who indicated needing help with ADLs or IADLs, there were follow-up items on the frequency that they received the help they needed. We dichotomized responses to always or almost always vs. sometimes, rarely, or never.

While we originally considered including a measure of camouflaging, that construct did not make the list of highest priority outcomes during the CBPR-nested Delphi process.^15^ As such, for this analysis, we use responses to a single item about need for masking as a marker for camouflaging. Specifically, our measure of community participation included the item: “When I participate in the community, I often feel like I **have to hide who I am** in order to be accepted.” Responses were on a 5-point Likert scale from strongly agree to strongly disagree.

We also included slightly adapted versions of the Everyday Experiences of Discrimination Scale^25^ and the Adverse Childhood Experiences scale.^26^

### Data Analysis

#### Structural validity and measurement invariance

We evaluated structural validity separately for the direct-report (DR), direct-report with support (DR-S), and caregiver-report (CR) groups. For the DR and DR-S groups, which had large enough samples, we randomly split the sample into two halves and conducted independent exploratory (EFA) and confirmatory factor analyses (CFA).^27^ For the CR group, we ran the EFA and CFA with the same sample, understanding that the sample size is limited and the results may not be robust and stable. We tested the measurement invariance of the burnout measure across the DR and DR-S groups before we decided to combine these two groups for other analyses.

#### A priori hypothesis testing for construct validity

We assessed construct validity via a priori hypothesis testing, using pair-wise correlations and t-tests. We converted t-scores to Cohen’s d’s to aid with assessing magnitude of correlations. In line with conceptual models of autistic burnout, we hypothesized that autistic burnout would have medium to strong positive correlations with a variety of stressors that might contribute to its development, such as experiences of discrimination, barriers to communication, and having experienced a major life event in the preceding 6 months. Conversely, we expected it to be negatively associated with various measures of support including social support, adequacy of supports and services, and receipt of help for basic or instrumental activities of daily living among those who felt they needed support for such activities.

Given widespread beliefs about the negative consequences of autistic burnout, we hypothesized medium to strong negative associations with broad life outcomes such as quality of life, flourishing, and overall health and wellbeing, as well as with social outcomes such as community participation and employment satisfaction. Finally, given what is known about the co-occurrence of autistic burnout with other health conditions, we hypothesized medium to strong positive associations between autistic burnout and depression, anxiety, and health interference.

#### Discriminant validity between autistic burnout and depression

To further evaluate longitudinal discriminant validity between autistic burnout and depression, we conducted item-level and scale-level analyses across three waves (T1– T3).^28^ First, we estimated longitudinal confirmatory factor analyses using item-level indicators to compare a hypothesized two-factor model, with separate autistic burnout and depression factors, to a one-factor general distress model. To do so, we used all 23 items from the AABM-R and the Patient Health Questionnaire (PHQ-9) together, as if they came from one scale. We then tested configural and metric invariance across waves to assess stability of the measurement structure over time. We specified residual correlations between identical items across waves to account for item-specific stability.

#### Preliminary criterion validity and score interpretation

We employed Receiver Operating Characteristic (ROC) analysis to assess preliminary criterion validity of the AABM-R (Bradley, 1997; Fawcett, 2006). As there is no established diagnostic gold standard for autistic burnout, we used a single item of perceived autistic burnout, “Do you think you are experiencing autistic burnout now?” The primary analysis compared participants responding Yes or No, excluding “I don’t know” responses. Two sensitivity analyses alternatively grouped “I don’t know” responses with No and then Yes.

The primary scoring specification used summed total scores among participants completing all 14 items. Sensitivity analyses used prorated scores for participants with incomplete item data. We compared mean item scores across criterion-response groups using the Kruskal–Wallis test followed by Holm-adjusted pairwise Wilcoxon rank-sum tests.

We estimated AUCs and 95% confidence intervals using 2,000 stratified bootstrap samples. Using the primary complete-item score and Yes-versus-No criterion, we evaluated candidate score ranges of 0–22, 23–32, and 33–56. For each range, we calculated the likelihood ratio as the probability of scoring in that range among participants responding Yes divided by the corresponding probability among participants responding No.^29^

## Results

### Structural Validity and Measurement Invariance

We found that the 14 items of autistic burnout were clearly loaded on a single-factor of Autistic Burnout in each of the separate EFAs using half the DR subsample, half the DR-support subsample, and the full CR sample. High item factor loadings (above .60), a high ratio of the first eigenvalue over the second eigenvalue (above 3), high Olkin measures (>= .899), and significant Bartlett’s test (*p* < .001) all supported the single-factor model.

In CFAs, the one-factor model of the autistic burnout measure had adequate fit for the second half of the DR subsample and second half of the DR-support subsample, with a Root Mean Square Error of Approximation (RMSEA) of .10 and .10, Comparative Fit Indices (CFI) of .93 and .93, and a Standardized Root Mean Square Residual (SRMR) of .04 and .05, respectively. In contrast, the one-factor model had inadequate fit for the CR sample, with a RMSEA of .20, CFI of .85, and SRMR of .07, potentially due to the small sample size (N = 66). **Tables 1 and 2** show the fit indices and standardized item factor loadings, respectively, from all 3 CFA models.

**Table 1:** Fit Indices from Confirmatory Factor Analysis (CFA) of AASPIRE Autistic Burnout Measure - Revised.

| Sample | $\chi^2$ | df | p | RMSEA [90% CI] | CFI | SRMR |
| --- | --- | --- | --- | --- | --- | --- |
|  |  |  | < |  |  |  |
| DR Subsample 2 | 297.39 | 73 | .001 | .102 [.090, .115] | .927 | .043 |
|  |  |  | < |  |  |  |
| DR-Support Subsample 2 | 141.95 | 74 | .001 | .100 [.075, .125] | .927 | .053 |
|  |  |  | < |  |  |  |
| CR – All | 251.12 | 73 | .001 | .197 [.170, .224] | .848 | .069 |

RMSEA = Root Mean Square Error of Approximation; CFI = Comparative Fit Index; SRMR = Standardized Root Mean Square Residual.

**Table 2:** Standardized Item Factor Loadings in Confirmatory Factor Analyses for Samples of Different Report Types.

| Items | Direct Report | Direct Report with Support | Caregiver Report |
| --- | --- | --- | --- |
| For at least the past three months... |  |  |  |
| I've been having more trouble **thinking** than I usually do.<br>For example, **more trouble** understanding things, making decisions, or solving problems. | .80 | .78 | .87 |
| I've been having more trouble than usual **controlling my emotions**.<br>For example, I have been moodier or more irritable than usual. | .69 | .83 | .78 |
| I've been having more problems than usual with **sensory sensitivities**.<br>For example, **more trouble** with sounds, lights, textures, colors, or smells. Trouble could be feeling bothered, not being able to ignore, or getting easily overstimulated. | .74 | .75 | .82 |
| I've been having more **extreme reactions to stress** than I usually do.<br>For example, **more meltdowns** or **shutdowns**. | .73 | .70 | .77 |
| I've been having a harder time than usual **controlling my actions**.<br>For example, **talking out of turn, screaming, pulling my hair, or throwing things more often**. | .62 | .61 | .74 |
| I've been having a harder time than usual **getting along with people**. | .72 | .77 | .74 |
| I've been having a harder time than usual **communicating**.<br>For example, **more trouble** finding the right words, speaking to people, or getting my point across. | .83 | .83 | .88 |
| I've been having a harder time than usual **taking care of myself**.<br>For example, **more trouble** eating, showering, cleaning, or shopping. | .81 | .68 | .90 |
| I've been having a harder time than usual handling **work, school, volunteering, or other regular activities**. | .85 | .72 | .88 |
| I've been having a harder time than usual **completing tasks**.<br>For example, **more trouble** getting started, figuring out the right steps, or doing the steps. | .83 | .84 | .93 |
| I have **wanted to stay away from other people** more often than I usually do.<br>For example, by avoiding social situations or staying home alone **more than usual**. | .72 | .72 | .79 |
| I've been **avoiding activities that require effort**, even if I like them, more often than I usually do. | .76 | .83 | .79 |
| I've been having a harder time than usual **remembering things**. | .73 | .70 | .85 |
| I've felt more **exhausted** than I usually do.<br>For example, feeling **extremely tired**. | .79 | .80 | .73 |

Given the inadequate fit of the CFA model for the CR sample, we only conducted measurement invariance analyses across the DR and DR-support subsamples, to establish the equivalence of the underlying autistic burnout construct across the two subsamples. We found that the measurement of the 14-item AABM-R demonstrated equivalence as equal residual variances (as well as configural and metric equivalence), but not equal intercepts (**Table 3**). That means the underlying meaning of autistic burnout and the importance of each item for capturing the idea of burnout was equivalent for the participants in the two subsamples (i.e., the meaning of their burnout scores appeared comparable), but the DR and DR-support groups may use the response scale of the autistic burnout measure somewhat differently.

**Table 3:** Measurement Invariance of AASPIRE Autistic Burnout Scale - Revised Across DR and DR-Support Samples.

| Model | $\chi^2$ (df) | RMSEA [90% CI] | CFI | SRMR | Comparison | $\Delta\chi^2$ (Δdf) | Significance |
| --- | --- | --- | --- | --- | --- | --- | --- |
| Configural | 671.99 (146) | .097 [.089, .104] | .932 | .041 | — | — | — |
| Metric | 687.26 (160) | .092 [.085, .100] | .932 | .049 | Metric vs. Configural | 15.27 (14) | $p > .05$ |
| Residual | 708.30 (174) | .089 [.082, .096] | .931 | .049 | Residual vs. Metric | 21.04 (14) | $p > .05$ |
| Intercept | 778.35 (188) | .090 [.084, .097] | .924 | .057 | Intercept vs.<br>Residual | 70.05 (14) | $p < .001$ |

The Configural model estimates factor loadings, residual variances, and intercepts freely. The Metric model constrains factor loadings to be equal. The Residual model constrains factor loadings and residual variances to be equal. The Intercept model constrains loadings, residual variances, and intercepts to be equal. $\Delta\chi^2$ values calculated from raw loglikelihood outputs.

Considering the inconclusive CFA results for the CR subsample, we decided to conduct the remainder of our reliability and validity analyses twice: primary analyses include the entire sample (N = 835), while secondary analyses exclude the CR subsample (N = 767). Secondary analyses did not significantly change any conclusions.

### A-priori hypothesis testing for construct validity

Autistic burnout was correlated, as hypothesized, in the expected directions, with all of the outcomes we tested, including quality of life (*r* = −0.47), overall health (*r* = −0.56), health interference (*r* = 0.59), flourishing (*r* = −0.47), depression (*r* =0.64), anxiety (*r* = 0.59), community participation (*r* = −0.50), barriers to communication (*r* = 0.43), overall employment satisfaction (*r* = −0.36), social support (*r* = −0.39), adequacy of supports and information (*r* = −0.46), barriers to healthcare (*r* = 0.40), experiences of discrimination (*r* = 0.44), and adverse childhood events (*r* = 0.41). Among people who needed support with either ADLs, IADLs, or both, autistic burnout was lower in those who received the support they needed (*d* = 0.94 for ADLs; *d* = 0.72 for IADLs). Finally, we found that burnout was higher in participants who had experienced a major life event in the past 6 months (*d* = 0.49), and in participants who agreed or strongly agreed to the item about masking (*d* = 0.88). *P*-values were <0.001 for all associations.

### Discriminant Validity with Depression

Longitudinal item-level CFA provided evidence that autistic burnout and depressive symptoms were related but distinct constructs. The hypothesized two-factor configural model (with a factor for depression and a second factor for autistic burnout) demonstrated acceptable fit, CFI = .909, TLI = .903, RMSEA = .042, SRMR = .050, and fit substantially better than a one-factor distress model that combined the two concepts (Δχ²(12) = 1654.5, *p* < .001; **Table 4**). Results supported metric invariance across T1– T3 (Δχ²(46) = 46.95, *p* = .434), indicating that item loadings were stable over time for both autistic burnout and depression separately. Latent correlations between autistic burnout and depression were moderate to large across waves, *r* = .68–.72, suggesting substantial association but not redundancy.^30,31^ Together, these findings support longitudinal discriminant validity of autistic burnout relative to depressive symptoms.

**Table 4:** Longitudinal Discriminant Validity Between Autistic Burnout and Depression: Confirmatory Factor Analysis Fit and Measurement Invariance.

| Model | $\chi^2$ | df | CFI | TLI | RMSEA [90% CI] | SRMR | Model comparison | $\Delta\chi^2$ (Δdf) | *p* |
| --- | --- | --- | --- | --- | --- | --- | --- | --- | --- |
| One-factor general distress model | 8,210.68 | 2,205 | .833 | .823 | .057 [.056, .058] | .091 | — | — | — |
| Two-factor configural model | 5,457.05 | 2,193 | .909 | .903 | .042 [.041, .043] | .050 | Configural vs. one-factor | 1,654.50 (12) | <.001 |
| Two-factor metric-invariance model | 5,524.56 | 2,239 | .909 | .904 | .042 [.040, .043] | .053 | Metric vs. configural | 46.95 (46) | .434 |

AABM-R = AASPIRE Autistic Burnout Measure–Revised; PHQ-9 = Patient Health Questionnaire–9; CFI = comparative fit index; TLI = Tucker–Lewis index; RMSEA = root mean square error of approximation; SRMR = standardized root mean square residual. Models were estimated using robust maximum likelihood with full-information maximum likelihood for missing data. $\chi^2$ values are robust scaled statistics; $\Delta\chi^2$ values are Satorra–Bentler scaled difference tests. The metric model constrained corresponding factor loadings to equality across waves.

### Criterion Validity and Scoring Interpretation

Among the 830 participants with a valid response to the current-burnout item, 224 responded “No,” 260 responded “Yes,” and 346 responded “I don’t know.” Mean AABM-R scores on the 0-4 metric differed across the No, I don’t know, and Yes groups (1.21, 1.81, and 2.64, respectively), with all Holm-adjusted pairwise Wilcoxon differences significant (at *p* < .001). In the primary ROC analysis (**Figure 1**), the AUC for distinguishing Yes-versus-No was .886 (95% bootstrap CI [.854, .915]). When ‘I don’t know’ responses were alternatively grouped with No or Yes, the AUCs were .808 (95% CI [.775, .839]) and .770 (95% CI [.735, .805]), respectively (N = 801). Results were essentially unchanged when scores were prorated to retain incomplete cases or caregiver reports were excluded.

**Figure 1.**
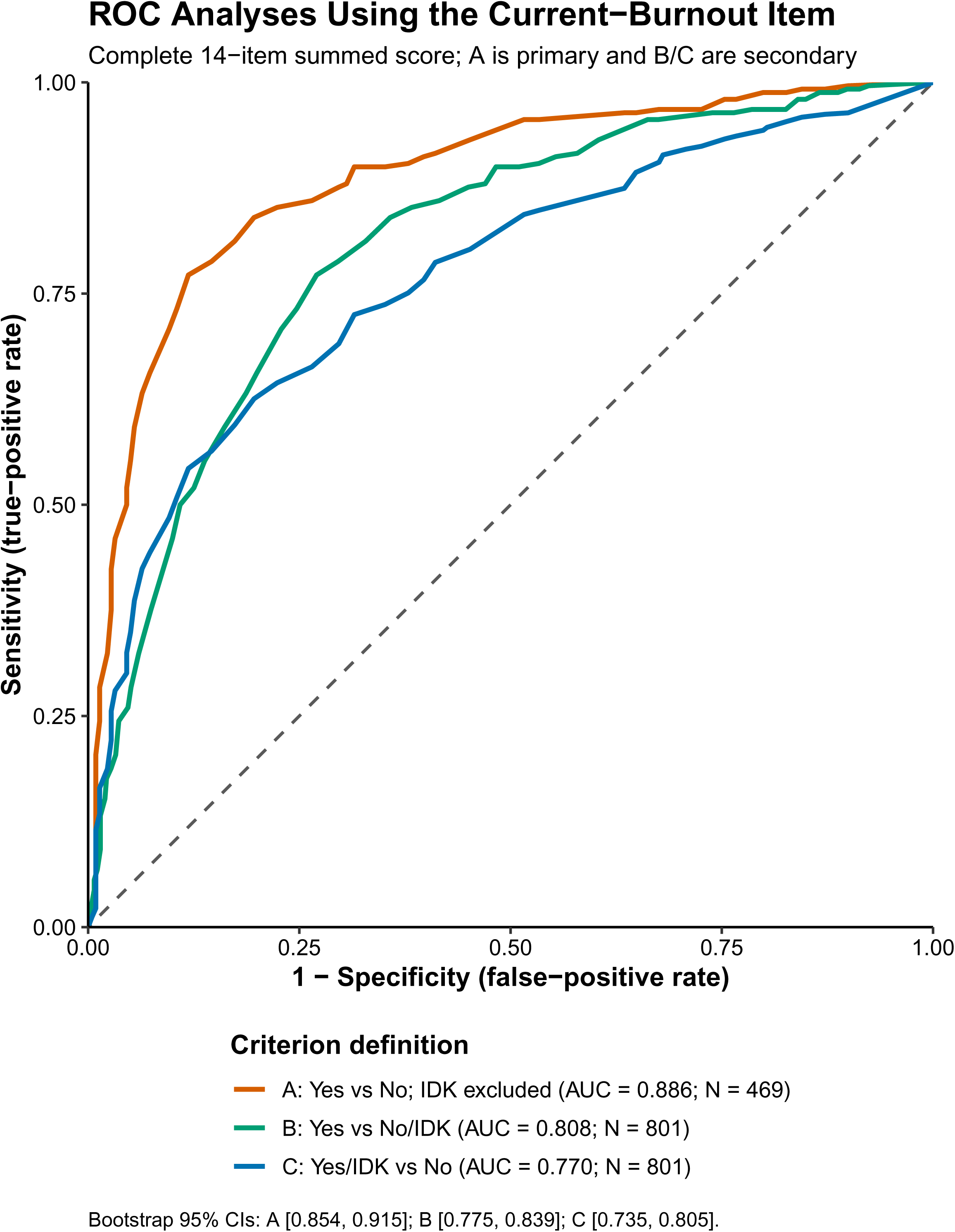
Receiver operator curve (ROC) for AASPIRE Autistic Burnout Scale – Revised compared to self-assessment of currently experiencing autistic burnout.

We evaluated two candidate thresholds defining three risk groups: 0-22 low probability; 23-32 unclear; and >=33 high probability of autistic burnout. A dichotomous threshold of 33 yielded sensitivity of .71 and a specificity of .90 while a dichotomous threshold of 23 yielded a sensitivity of .90 and a specificity of .68. Multi-level likelihood ratios supported a three-level scoring approach. Scores of 0–22 decreased the likelihood of self-reported current autistic burnout (LR = 0.15, 95% CI [0.09, 0.21]); scores of 23–32 provided little change in likelihood (LR = 0.88, 95% CI [0.61, 1.25]); scores of 33–56 increased the likelihood of self-reported current autistic burnout (LR = 7.38, 95% CI [5.09, 12.00]).

## Discussion

While multiple groups have provided promising psychometric evidence for the pre-publication version of the AABM, our own team members and outside experts with lived and professional experience raised concerns about the accessibility of the instrument for people with co-occurring intellectual disability or higher support needs. They also requested a briefer measure that could be used more easily in real-world service or clinical settings. As such, we revised and shortened the original AABM, creating the 14-item AABM-R, and we tested it as part of a larger longitudinal study with a heterogeneous population of over 800 autistic adults recruited from healthcare systems, disability service systems, and the community. In an overview paper discussing initial psychometric analyses of all 19 outcome measures in the AASPIRE Measurement Toolkit, we recently reported that the direct report version of the AABM-R has strong content validity, internal consistency reliability, 2-week test-retest reliability, convergent validity, and 6-month responsiveness to change.^18^ In the current paper, we further tested the AABM-R’s psychometric properties via a series of more in-depth analyses focused on structural validity, measurement invariance, construct validity, and clinically meaningful cut-point scoring, as well as testing differentiations between autistic burnout and depression.

Previous structural analyses of the original AABM had mixed results, with some studies finding it largely fit into a single factor,^4,5^ while another proposed a 4-factor model.^6^ We found that the shorter AABM-R constitutes a clear single factor, at least for participants completing it directly, with or without support. Results were less clear for those using the caregiver report version of the survey, perhaps due to the small number of proxy reporters or because proxies cannot reliably report on another person’s experience of burnout.

To our knowledge, no previous studies of autistic burnout instruments looked at measurement invariance for people who completed surveys with varying levels of support, perhaps in part because most prior studies included internet-based convenience samples who primarily could take part in survey studies independently.^2^ Our sample included a substantive number of participants (N=182) who required support to take part in the study directly. We found evidence of measurement invariance between those who participated with vs. without support, suggesting that the direct report version of the AABM-R functions similarly for participants with higher support needs. Similarly, our study extends the literature by demonstrating that autistic burnout is a prevalent, measurable construct, even in more heterogeneous populations with a broader range of communication challenges, age at diagnosis, living arrangements, or ADLs/IADLs support needs.

We assessed the scale’s construct validity using a priori hypothesis testing. We based our hypotheses on the growing conceptualization of autistic burnout as an imbalance of life stressors and supports, as well as the observation that burnout may lead to worsened health, wellbeing, and quality of life.^1,2,11^ Data supported all of our *a priori* hypotheses, finding associations in the expected directions with stressors such as major life events, barriers to communication, experiences of discrimination, masking, and early childhood adverse experiences; and supports and services, including social support, adequacy of support and information, receipt of needed support for ADLs or IADLS, and barriers to healthcare. We also found associations, in the expected directions, with other health issues such as depression, anxiety, and health interference, and with broad life outcomes such as quality of life; flourishing; and overall physical, mental, and social wellbeing. These findings both add evidence of the scale’s construct validity, and they support the growing consensus on the nature and importance of autistic burnout.

The autistic community has always asserted that autistic burnout is distinct from depression.^1,7^ However, in our experience discussing the concept of autistic burnout in academic and clinical circles, critics commonly contest the distinction based on their presuppositions. While evidence to support the distinction is growing,^2,5^ the creators of the ABSI questioned the ability of the original AABM to distinguish between autistic burnout and depression.^4^ Our study provides more rigorous evidence that autistic burnout and depression are related, but distinct, and that the AABM-R can distinguish between the two constructs despite their high co-occurrence and the overlap in some of their features (e.g., fatigue, diminished concentration). Specifically, a two-factor model separating autistic burnout and depression fit substantially better than a one-factor general distress model. Moreover, metric invariance indicated that the item loadings supporting the distinction were stable across waves.

A strength of both the AABM and the AABM-R is that, unlike the ABSI, they do not assume that the participant already recognizes that they have ever had autistic burnout. As such, they have the potential to be used as a screening tool in people who may or may not have ever experienced autistic burnout or may or may not even be aware of the concept. Past studies of both the AABM and the ABSI were conducted in populations where all or almost all participants had experienced burnout.^3,4^ While there is no clinical gold standard for the diagnosis of autistic burnout at this time, our ROC analysis, comparing AABM-R scores to participant’s self-assessment of being in an episode of autistic burnout, allowed us to offer preliminary clinical cut-offs for low, medium, and high probability of burnout. Such cut-offs are not diagnostic but may encourage clinicians to further assess for autistic burnout with a more in-depth clinical interview.

Our study has several limitations. We put considerable effort, using our CBPR process, into making all aspects of the study as accessible as possible.^17^ Our success in this goal likely allowed many participants who would have needed to take the study directly with support to take it independently, and many participants who would have taken part via proxy reporter to take surveys directly with support. As such, we did not have a large enough sample of participants who completed the survey using the caregiver report version to confirm the single factor model for this group or assess measurement invariance. While initial psychometric analyses provided some evidence supporting the caregiver report version’s internal consistency reliability and construct validity,^18^ we would recommend additional testing of the caregiver report version of the measure, with a larger sample size of caregiver reporters, to understand its structural validity.

While we consider camouflaging to be a common and important stressor that can lead to burnout,^1^ other researchers have placed a more central role on camouflaging and have criticized the original AABM for not having a very strong association with camouflaging, at least as measured by the Camouflaging Autistic Traits Questionnaire (CAT-Q).^4^ The current study did not include a full measure for camouflaging because the construct did not make it into the group of highest priority outcomes during our CBPR-nested Delphi process.^15^ Moreover, team members felt that common measures of camouflaging such as the CAT-Q do not adequately distinguish between 1) wanting to or feeling the need to camouflage, 2) using camouflaging behaviors, 3) no longer being able to camouflage due to factors such as burnout, or 4) never having been able to effectively camouflage due to issues such as being non-speaking or having a co-occurring intellectual disability (also see ^32^). We found that agreeing with a single item about feeling the need to mask was strongly associated with higher autistic burnout scores. However, more research is needed to learn how to adequately measure camouflaging without confusing need, intent, actions, exhaustion, and ability, and to understand its relationship with autistic burnout, especially in heterogeneous populations of autistic adults.

It is theoretically possible that what is currently termed “autistic burnout” might more accurately be understood as increased susceptibility to burnout or differing presentations of burnout among autistic people, rather than a wholly separate construct than the forms of professional or caregiver burnout that have been described in psychological research.^33^ Even though other research has directly compared the original AABM to traditional measures of burnout, and has suggested that the AABM may be somewhat better at identifying individuals experiencing autistic burnout,^5^ we did not include a traditional measure of burnout, so we cannot directly add to the literature distinguishing autistic burnout from professional or caregiver burnout.

This paper reports on univariate associations between AABM-R scores and potential contributors to or consequences of autistic burnout simply to assess construct validity. While such associations confirm current conceptualizations of autistic burnout, we are actively conducting more detailed multivariate longitudinal analyses to understand the temporality between potential causes and consequences of autistic burnout and plan to publish those separately.

The naming of “autistic burnout” may unfortunately be misleading. Some authors have proposed reframing autistic burnout as “autistic exhaustion” to emphasize depletion rather than workplace connotations,^34^ while others have suggested that burnout may represent part of a broader “autistic exhaustion syndrome” encompassing inertia and shutdown and potentially being linked to autistic catatonia.^4^ Autistic burnout may also be a part of a more general “disability burnout” experienced by people with many different types of disabilities. Finally, panelists at a recent round table discussion on burnout, long COVID, and myalgic encephalomyelitis / chronic fatigue syndrome (ME/CFS) in autism and ADHD suggested that the symptoms of autistic or ADHD burnout may be the common manifestation of any “chronic energy depletion syndrome” in neurodivergent patients (Grillo et al, in preparation). As such, the term “burnout” may inappropriately denote a psychological etiology when in fact autistic burnout symptoms may be due to chronic mitochondrial dysfunction or other physiologic etiologies.

Similarly, research has shown an association between autism and central sensitivity syndromes.^35,36^ It is theoretically possible that autistic burnout may contribute to that association, especially in regard to sensory hypersensitivities. While we prefer the chronic energy depletion conceptualization, we have chosen to at least temporarily continue to use the term “autistic burnout” given that that is what the autistic community currently calls it. It is possible that the measure may need to be renamed in the future or may need to be adapted to more fully cover other forms of chronic energy depletion in broader neurodivergent populations.

## Conclusion

The AASPIRE Autistic Burnout Measure-Revised is a brief, accessible, easy-to-use instrument intended for use with heterogeneous populations of autistic adults in clinical, services, and/or research settings. Prior analyses from this data set support its internal consistency reliability, 2-week test-retest reliability, content validity, construct validity, and 6-month responsiveness to change. The AABM-R measures a construct that is empirically distinct from depression symptoms in cross-sectional and longitudinal measurement models. The direct report version has strong structural validity, with a single factor, and measurement invariance between those that take it with or without support. More research is needed about the caregiver report version of the scale. Based on ROC analyses, in comparison to participant’s self-assessment of currently being in a chronic episode of autistic burnout, we recommend provisional cut-offs of scores of 0-22 for decreased (LR= 0.15) and 33-56 for increased likelihood of chronic autistic burnout (LR = 7.47).

The longitudinal data from the AASPIRE Outcomes Project will allow us and others to better understand the potential contributors to and outcomes of autistic burnout. Use of this instrument will also help researchers better understand the effectiveness of interventions to prevent or treat autistic burnout in real world settings.

## Acknowledgements

We wish to acknowledge the great contributions, throughout the project, of the rest of the AASPIRE Outcomes Project team, including Shannon des Roches Rosa, Todd Edwards, Emanuel Frowner, Willi Horner-Johnson, Andrea Joyce, Clarissa Kripke, Julie Lounds-Taylor, Julia Love, Joelle Maslak, Katherine McDonald, Zack Siddeek, Ivanova Smith, Anna Furra Wallington, and Finn Gardner. We also wish to thank our research staff, including Joseph Vera, KJ Flores, Grace Herbert, Rachel Schuck, Janea Jones, Madeline Proctor, Reilly Caldwell, and Sullivan Swift, for help with recruitment, data collection and management, and regulatory compliance. Finally, we are grateful to the many community members and colleagues who helped with recruitment and to all the study participants for their time and openness.

## Author Contributions

Christina Nicolaidis: Conceptualization, formal analysis, funding acquisition, methodology, supervision, writing – original draft

Liu-Qin Yang: Conceptualization, formal analysis, funding acquisition, methodology, writing – original draft

Mathew C. Uretsky: Conceptualization, formal analysis, funding acquisition, methodology, writing – original draft

Dora Raymaker: Conceptualization, funding acquisition, methodology, writing – review and editing

Mary Baker-Ericzén: Conceptualization, funding acquisition, methodology, resources, supervision, writing – review and editing

Vivian Darlene Grillo: Conceptualization. writing – original draft

Steven K. Kapp: Conceptualization, methodology, writing – review and editing

Rachel Kripke-Ludwig: Conceptualization, methodology, writing – review and editing

Joelle Maslak: Conceptualization, methodology, writing – review and editing

Ian Moura: Conceptualization, methodology, writing – review and editing

Mirah Scharer: Investigation, methodology, project administration, supervision, writing – review and editing

Anna Wallington: Conceptualization, methodology, writing – review and editing

## Statements and Declarations

### Ethical considerations

The Institutional Review Board (IRB) at Portland State University (PSU) approved the project and served as the Single IRB (approval # 227674-18); Oregon Health & Science University, Vanderbilt University Medical Center, and San Diego State University waived authorization to the PSU IRB. Involvement by the Multnomah County Developmental Disability Services was subsumed under the PSU IRB. Participants or their legally authorized representatives gave written consent online or in-person.

### Consent for publication

Not applicable

### Declaration of conflicting interest

Christina Nicolaidis is Editor-in-Chief of *Autism in Adulthood*. Authors do not have any other conflicting interests to declare.

### Funding statement

This project was funded by the National Institute of Mental Health through Grant Award Number R01MH121407. It was also supported by the National Center for Advancing Translational Sciences (NCATS), National Institutes of Health, through Grant Award Number UL1TR002369. The content is solely the responsibility of the authors and does not necessarily represent the official views of the NIH.

### Data availability

All data from this study will be available through the National Institutes of Health (NIH) National Data Archive (NDA).

## Supplemental Materials

**Supplement A:** Full direct report and caregiver report versions of the AABM-R, including detailed comparison with original AABM.

**Supplement B:** Participant Characteristics

## AASPIRE Autistic Burnout Scale – Revised: Overview

### Description

This instrument identifies and measures chronic autistic burnout.

### Development

The [AASPIRE Burnout Project](https://aaspire.org/projects/mental-health/pilot-study-on-autistic-burnout-and-suicidal-behavior/) team used a community based participatory research (CBPR) approach to create the original version of AASPIRE Autistic Burnout Scale in 2019. Items in the original Autistic Burnout Scale are based on the results of their qualitative study about autistic burnout (<u>Raymaker et al., 2020</u>).

We are currently recommending use of the revised version of the Autistic Burnout Scale, which was revised for inclusion in the [AASPIRE Outcomes Project](https://aaspire.org/projects/outcomes/). The Outcomes Project team is using a CBPR approach to 1) create and validate the AASPIRE Measurement Toolkit; and 2) understand what predicts changes in outcomes over time (Nicolaidis et al., 2026a).

Revisions to the AASPIRE Burnout Scale included:

- Shortening the instrument
- Adding text to clarify the time frame
- Making minor changes to increase accessibility
- Adding detailed descriptions and vignettes to more clearly define concepts, and
- Adding items about the onset of symptoms

Each instrument in the [AASPIRE Measurement Toolkit](https://aaspire.org/measurement-toolkit) has self-and caregiver-reported versions. AASPIRE developed and tested the instruments with and for autistic adults with or without intellectual disability, but they could also be used with general populations or other people who may need cognitively accessible survey instruments.

The AASPIRE Outcomes Project community-academic team collaboratively revised this instrument for inclusion in the AASPIRE Measurement Toolkit.

See below for a <u>more detailed comparison between the original and adapted instrument</u>.

### Scoring

Each item on the Symptom Severity section of the Autistic Burnout Scale is scored on a 5-point Likert scale, from 0 (Strongly Disagree) to 4 (Strongly Agree). The total score is the sum of items 1-14. Scores can range from 0 to 56, with higher scores indicating more severe burnout. Additional items are intended for clinical or research purposes and are not part of the total score. Tentatively, we recommend using the following cut-offs for clinically significant burnout: 0-22 low probability; 23-32 intermediate probability; 33 and above: high probability. These interpretations should always be combined with clinical discretion.

### Psychometric Properties

The original AASPIRE Autistic Burnout Scale showed strong psychometric properties, though it was tested primarily with samples recruited from the internet (Bougoure et al., 2025; Mantzalas et al., 2024).

The revised version is being psychometrically tested as part of the AASPIRE Outcomes Study, a longitudinal cohort study following a heterogeneous sample of ∼850 autistic adults over time (Nicolaidis et al., 2026a).

Cognitive interview data demonstrated strong content validity. Survey data have demonstrated excellent <u>internal consistency reliability</u> (alpha=0.95) and <u>2-week test-retest reliability</u> (intraclass coefficient of 0.92), strong <u>convergent and discriminant validity</u>, and good <u>6-month responsiveness to change</u> (Nicolaidis et al., 2026b)

Preliminary analyses support that the scale has a single factor and good construct validity (Nicolaidis et al., 2026a).

Additional results about the scale’s structural and construct validity are pending publication and will be shared on the [AASPIRE Measurement Toolkit](https://aaspire.org/measurement-toolkit) site when available.

### Conditions for Use

The AASPIRE Autistic Burnout Scale – Revised is available for use with attribution in noncommercial, clinical, and research settings. Permission is required for instrument adaptation, modification, and for-profit use. For permissions inquiries, contact Christina Nicolaidis and AASPIRE at

## AASPIRE Autistic Burnout Scale – Revised: Direct Report Version

### Introduction

The following set of questions focuses on **big, negative, long-term changes** in your life.

- We are interested in things that you have been experiencing for **three months or longer.**
- Compare what you have been experiencing for three months or more to what is **usual for you**.
- If you have been feeling bad for a long time, think back to what things were like before you started feeling bad.

You can use the following links to get information on:

- <u>What it means to have been experiencing something for three months or longer</u>
- <u>How to compare experiences to what is usual for you</u>
- <u>Examples of how different people may answer these questions</u>

### Symptom Severity

Please mark how strongly you agree or disagree with each statement.

Please choose one answer per row.

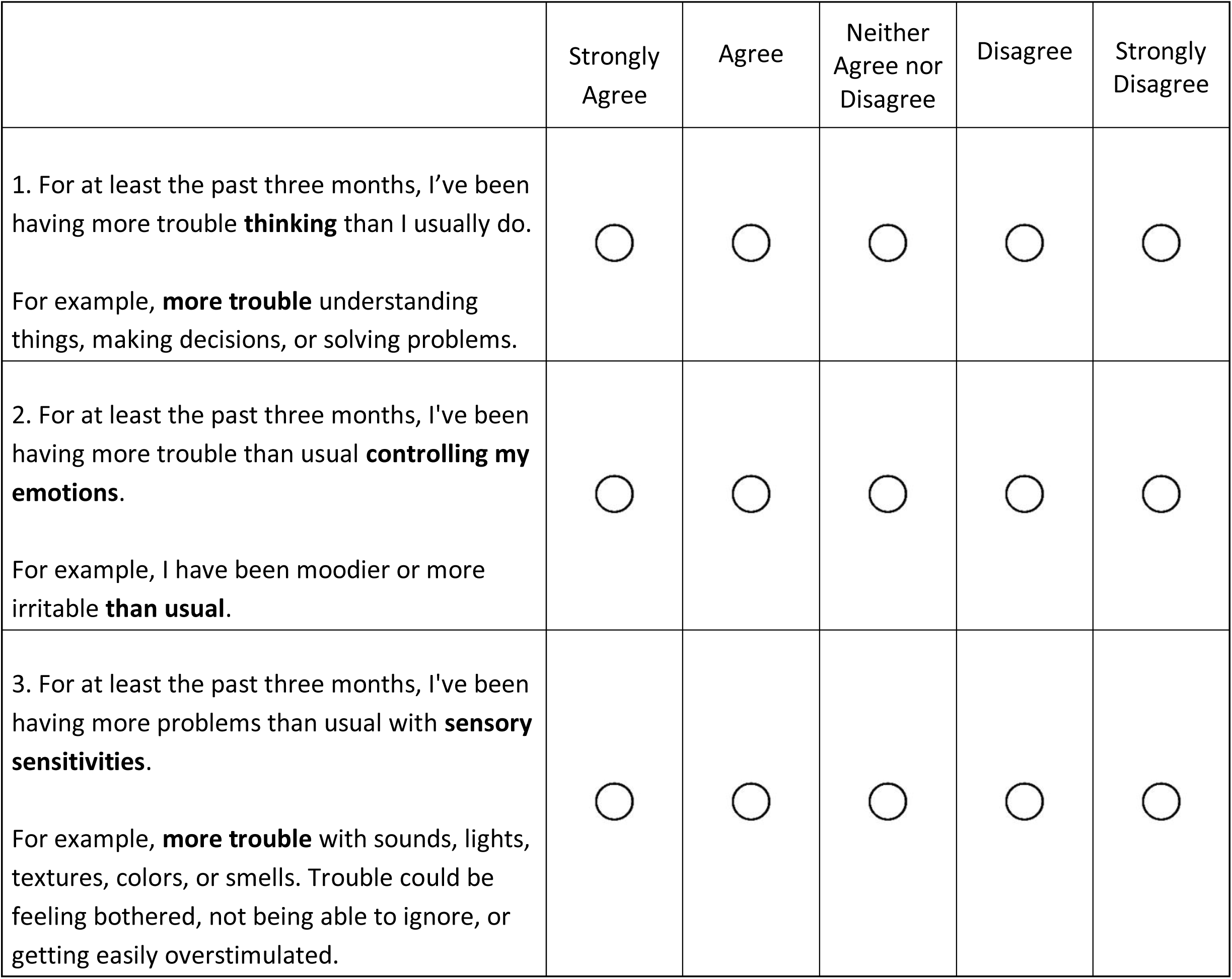
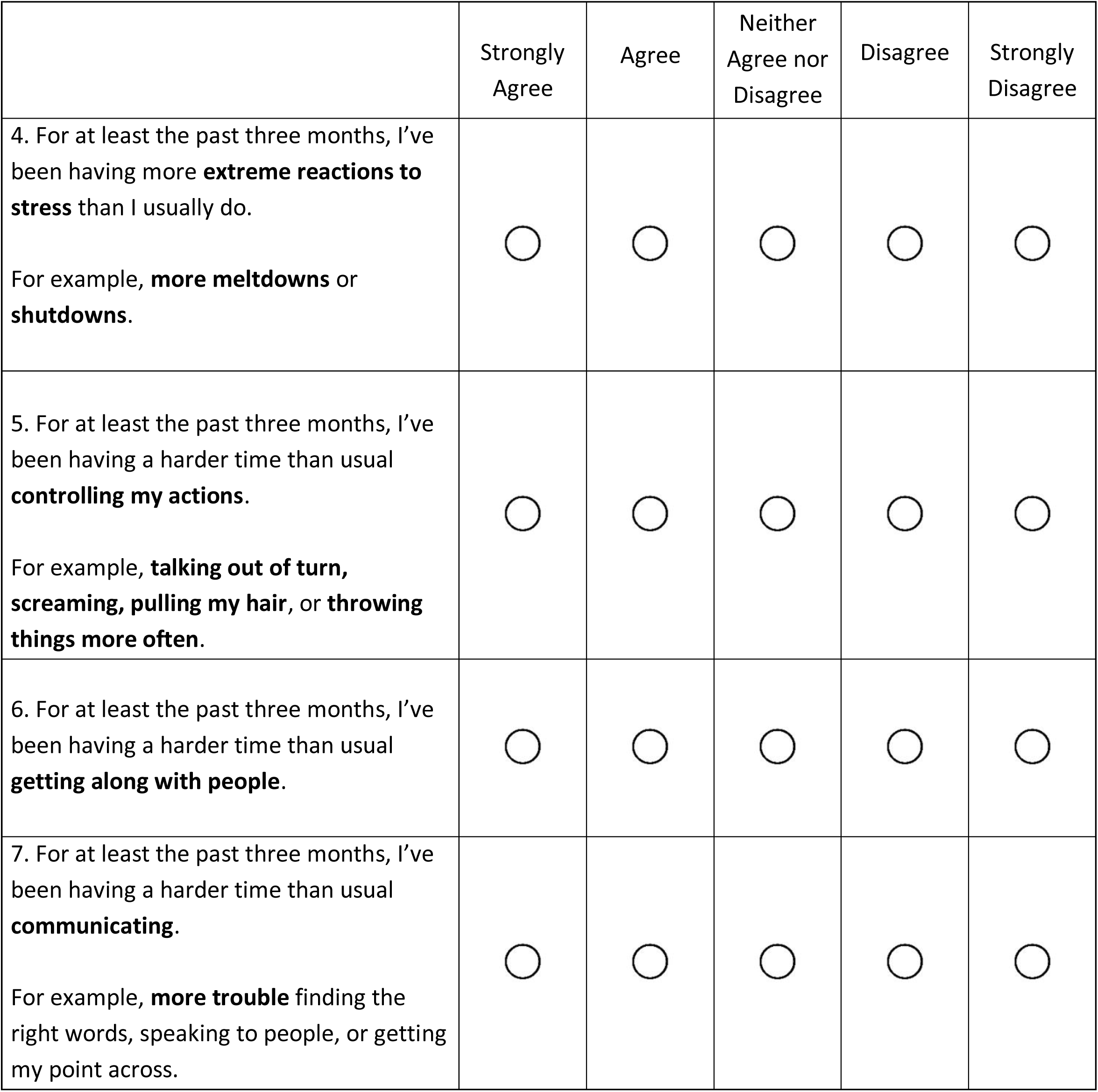
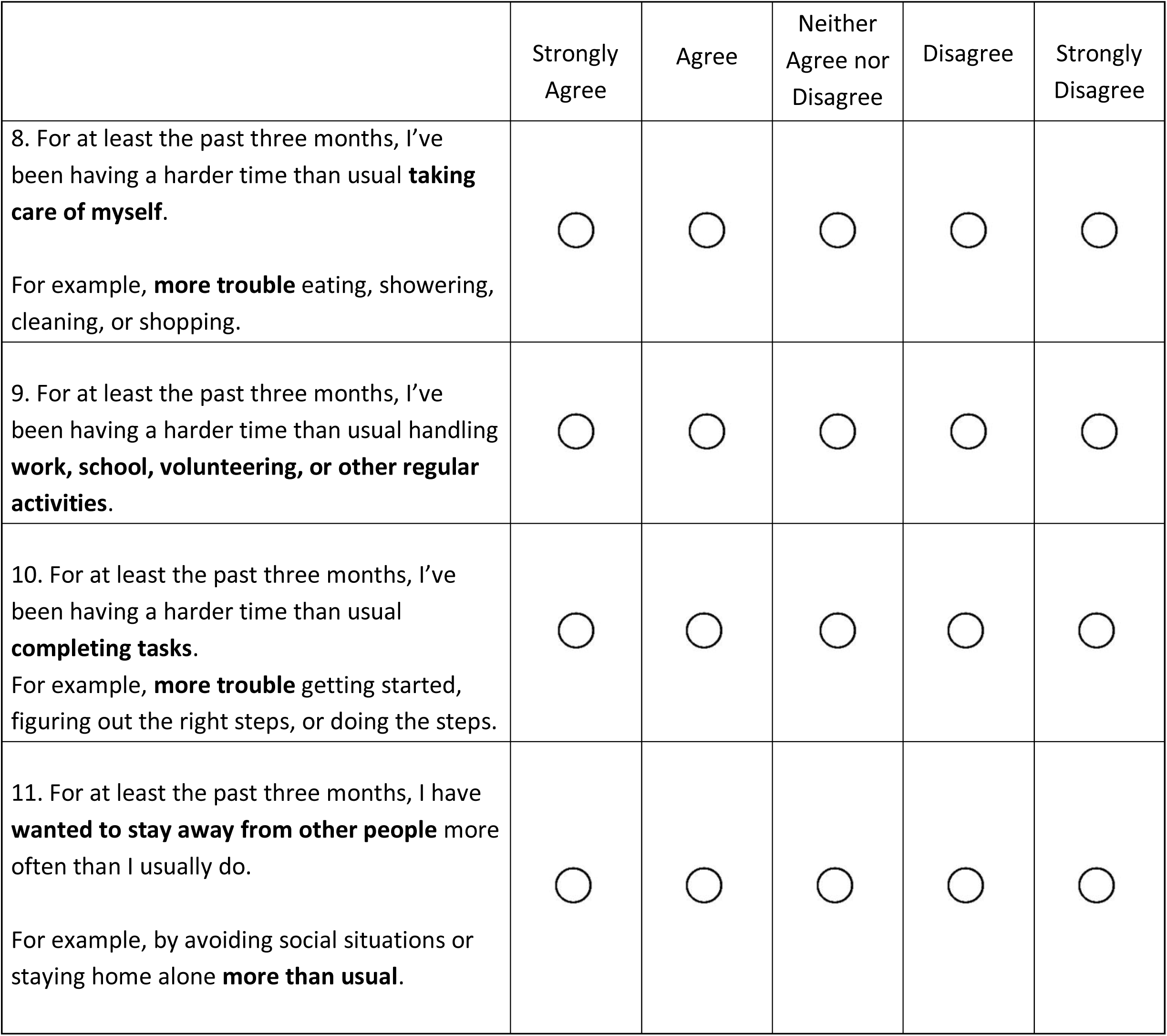
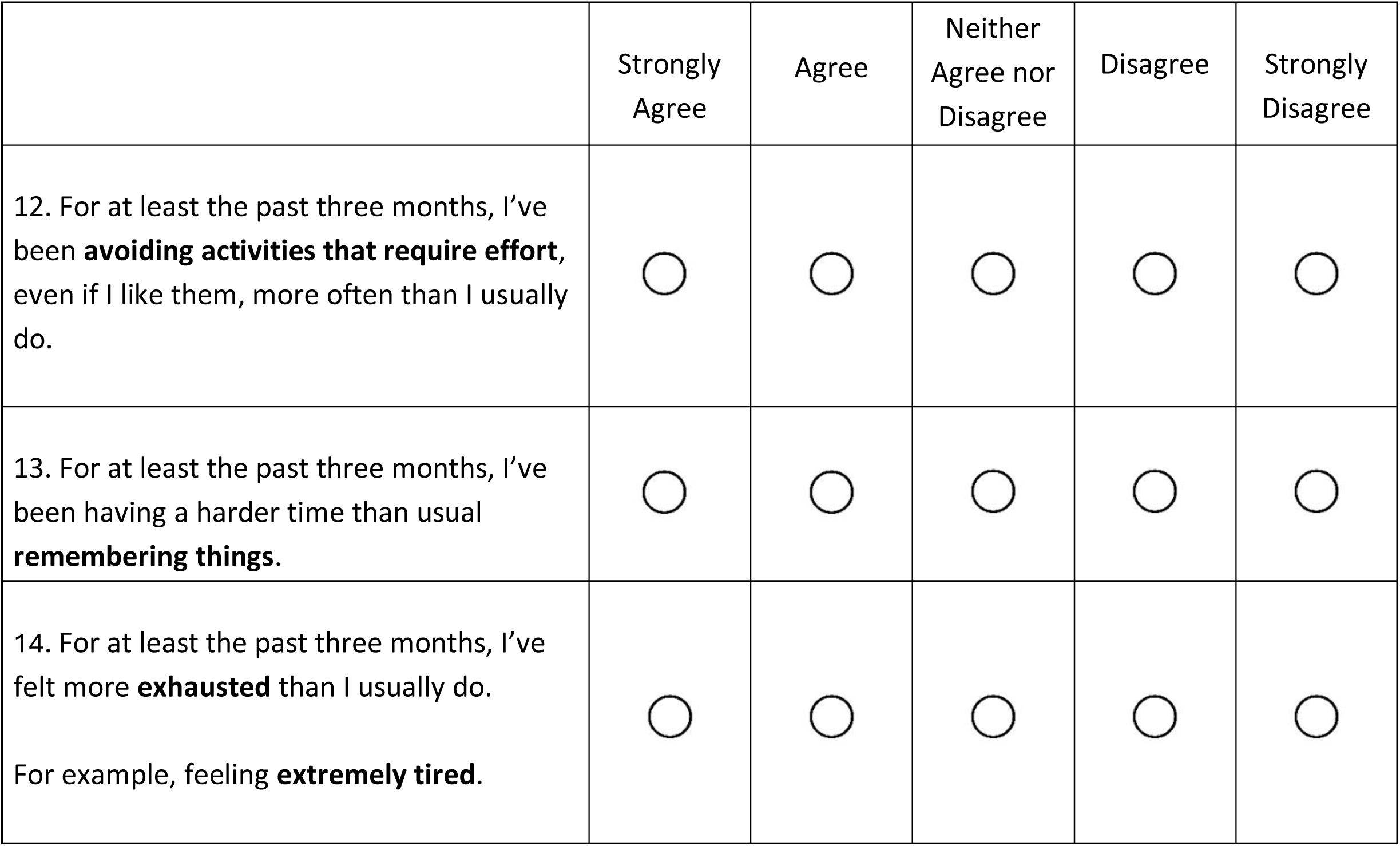

|  | Strongly Agree | Agree | Neither Agree nor Disagree | Disagree | Strongly Disagree |
| --- | --- | --- | --- | --- | --- |
| 1. For at least the past three months, I've been having more trouble **thinking** than I usually do.<br>For example, **more trouble** understanding things, making decisions, or solving problems. | ☐ | ☐ | ☐ | ☐ | ☐ |
| 2. For at least the past three months, I've been having more trouble than usual **controlling my emotions**.<br>For example, I have been moodier or more irritable **than usual**. | ☐ | ☐ | ☐ | ☐ | ☐ |
| 3. For at least the past three months, I've been having more problems than usual with **sensory sensitivities**.<br>For example, **more trouble** with sounds, lights, textures, colors, or smells. Trouble could be feeling bothered, not being able to ignore, or getting easily overstimulated. | ☐ | ☐ | ☐ | ☐ | ☐ |
| 4. For at least the past three months, I've been having more **extreme reactions to stress** than I usually do.<br>For example, **more meltdowns** or **shutdowns**. | ☐ | ☐ | ☐ | ☐ | ☐ |
| 5. For at least the past three months, I've been having a harder time than usual **controlling my actions**.<br>For example, **talking out of turn**, **screaming**, **pulling my hair**, or **throwing things more often**. | ☐ | ☐ | ☐ | ☐ | ☐ |
| 6. For at least the past three months, I've been having a harder time than usual **getting along with people**. | ☐ | ☐ | ☐ | ☐ | ☐ |
| 7. For at least the past three months, I've been having a harder time than usual **communicating**.<br>For example, **more trouble** finding the right words, speaking to people, or getting my point across. | ☐ | ☐ | ☐ | ☐ | ☐ |
| 8. For at least the past three months, I've been having a harder time than usual **taking care of myself**.<br>For example, **more trouble** eating, showering, cleaning, or shopping. | ☐ | ☐ | ☐ | ☐ | ☐ |
| 9. For at least the past three months, I've been having a harder time than usual handling **work, school, volunteering, or other regular activities**. | ☐ | ☐ | ☐ | ☐ | ☐ |
| 10. For at least the past three months, I've been having a harder time than usual **completing tasks**.<br>For example, **more trouble** getting started, figuring out the right steps, or doing the steps. | ☐ | ☐ | ☐ | ☐ | ☐ |
| 11. For at least the past three months, I have **wanted to stay away from other people** more often than I usually do.<br>For example, by avoiding social situations or staying home alone **more than usual**. | ☐ | ☐ | ☐ | ☐ | ☐ |
| 12. For at least the past three months, I've been **avoiding activities that require effort**, even if I like them, more often than I usually do. | ☐ | ☐ | ☐ | ☐ | ☐ |
| 13. For at least the past three months, I've been having a harder time than usual **remembering things**. | ☐ | ☐ | ☐ | ☐ | ☐ |
| 14. For at least the past three months, I've felt more **exhausted** than I usually do.<br>For example, feeling **extremely tired**. | ☐ | ☐ | ☐ | ☐ | ☐ |

### Additional Items

*The following items were also included in the Outcomes Project section on autistic burnout:*

#### Onset

*<If ‘Agree’ or ‘Strongly Agree’ options are selected for one or more items, Question **#15** and Question **#16** are displayed>*

15. We are sorry to hear that you are experiencing some of the changes described above.

**How quickly** did these changes seem to come on?

◦ It feels things got worse **within days or weeks** (for example, days or a few weeks after an illness or major life event, whether the event was good or bad).
◦ They **slowly built up over many months or years**.
◦ Other *<write-in>*

16. What do you think **may have brought on** the changes described in this section? (Check all that apply)

❑ **physical illness, surgery, or accident** <u>(click to see examples)</u>.
❑ **major life event or transition**, whether it was a good or bad thing <u>(click to see examples)</u>.
❑ **long-term mismatch** between expectations and your abilities, resources, and supports <u>(click to see examples)</u>.
❑ Other <*write-in*>

#### Self-Reflection

*<**Additional items** Question **#17** – Question **#19** included in survey are not a part of the scale>*

Many autistic people have talked about something called “autistic burnout.”

17. Before taking this survey, had you ever heard of “autistic burnout”?

◦ Yes, and I feel like I have a good idea of what it is
◦ Yes, but I don’t know much about it
◦ No

18. Do you think you have **ever** experienced autistic burnout?

◦ Yes
◦ No
◦ I do not know

19. Do you think you are experiencing autistic burnout **now**?

◦ Yes
◦ No
◦ I do not know

#### Standard Direct Report Items on Perceived Change and Use of Support to Participate

Overall, in the last six months, did the things this survey asked about…

◦ Get worse
◦ Stay about the same
◦ Get better

Did you get any help while answering the questions in this section of the survey?

◦ Yes
◦ No

*<If ‘Yes’ to get help question>*

What kind of help did you get while answering the questions in this section of the survey? (Check all that apply.)

❑ Someone helped me use a computer, smartphone, or other device (for example, they clicked on the answers I chose).
❑ Someone read the survey to me.
❑ Someone helped me understand what the questions or answers options meant.
❑ Someone helped me decide how to answer the questions on the survey.
❑ I got some other type of help to answer the questions.
❑ Someone answered the questions for me, without my input.

### Hyperlinks

#### Information on what it means for something to have been going on for at least 3 months

The questions ask about experiences that have been going on for **at least three months**:

- These things could have been going on for **3 months** or they could have been going on for **years.**
- It’s okay if you don’t remember exactly how long they have been going on.
- They don’t have to happen every single day for you to include them.
- Don’t include things that have only been going on for a few days or weeks.

#### How to compare experiences to what is “usual for you”

The questions ask you to compare these experiences to **what is “usual” for you**.

- “Usual” means whatever you consider **normal for you**, or to be most **typical for you**. This may be your current experience, or it may not be.
- If you have a lot of variation in your experiences with some of these things, that is okay. Think about whether there’s a difference between the current variation and the usual variation.

#### Examples of how different people may answer the questions

- Loud sounds have always bothered Maria, but in the past, she could usually cope with it.

◦ Now it’s a lot worse. Almost any noise feels really painful.
◦ She doesn’t remember exactly when she started having more trouble tolerating sounds, but she thinks it’s been gradually getting worse since she started a stressful new job about a year ago.
◦ She should answer “Strongly Agree” to the question about having more problems than usual with sensory sensitivities.
- Sam has always had a hard time controlling their emotions.

◦ There hasn’t been any change in their ability to control their emotions.
◦ Sam should answer “Strongly Disagree” to the question about having more trouble than usual controlling emotions because the trouble hasn’t changed.
- Jamal usually has some trouble concentrating.

◦ Jamal just got some bad news. He is having a much harder time concentrating today than he usually does.
◦ Before getting the bad news, Jamal was concentrating about as well as he normally does.
◦ Jamal should answer “Strongly Disagree” to the question about having more trouble concentrating because the change hasn’t been going on for at least three months.

#### Examples of a physical illness, surgery, or accident

- Getting COVID or another infection
- Having a stroke, getting really sick, being hospitalized, or having major surgery
- Getting hurt in an accident

#### Examples of a major life event or transition

- Starting, graduating, or leaving a school, college, or training program
- Moving to a new home or living with different people
- Getting into a new romantic relationship, getting married, or starting a family
- Breaking up with someone or getting divorced
- Getting new caregivers or supporters
- A change in your job or employment
- Being in a really scary or dangerous situation
- The death of a loved one or the loss of a supporter

#### Examples of a long-term mismatch between expectations and your abilities, resources, and supports

◦ Trying too hard, for too long, to meet your own or other people’s expectations, without getting the support or accommodations you would need to be successful
◦ Working too long without a break

## AASPIRE Autistic Burnout Scale – Revised: Caregiver Report Version

### Introduction

The following set of questions will focus on **big, negative, long-term changes** in their life.

- We are interested in things that you think they have been experiencing for **three months or longer.**
- Compare what you think they have been experiencing for three months or more to what you think is **usual for them**.
- If you think they have been feeling bad for a long time, think back to what things were like before they started feeling bad.

<u>You can use the following links to get information on:</u>

- <u>What it means to have been experiencing something for three months or longer</u>
- <u>How to compare experiences to what is “usual” for them</u>
- <u>Examples of how different people may answer these questions</u>

### Symptom Severity

Please mark how strongly you agree or disagree with each statement.

Please choose one answer per row.

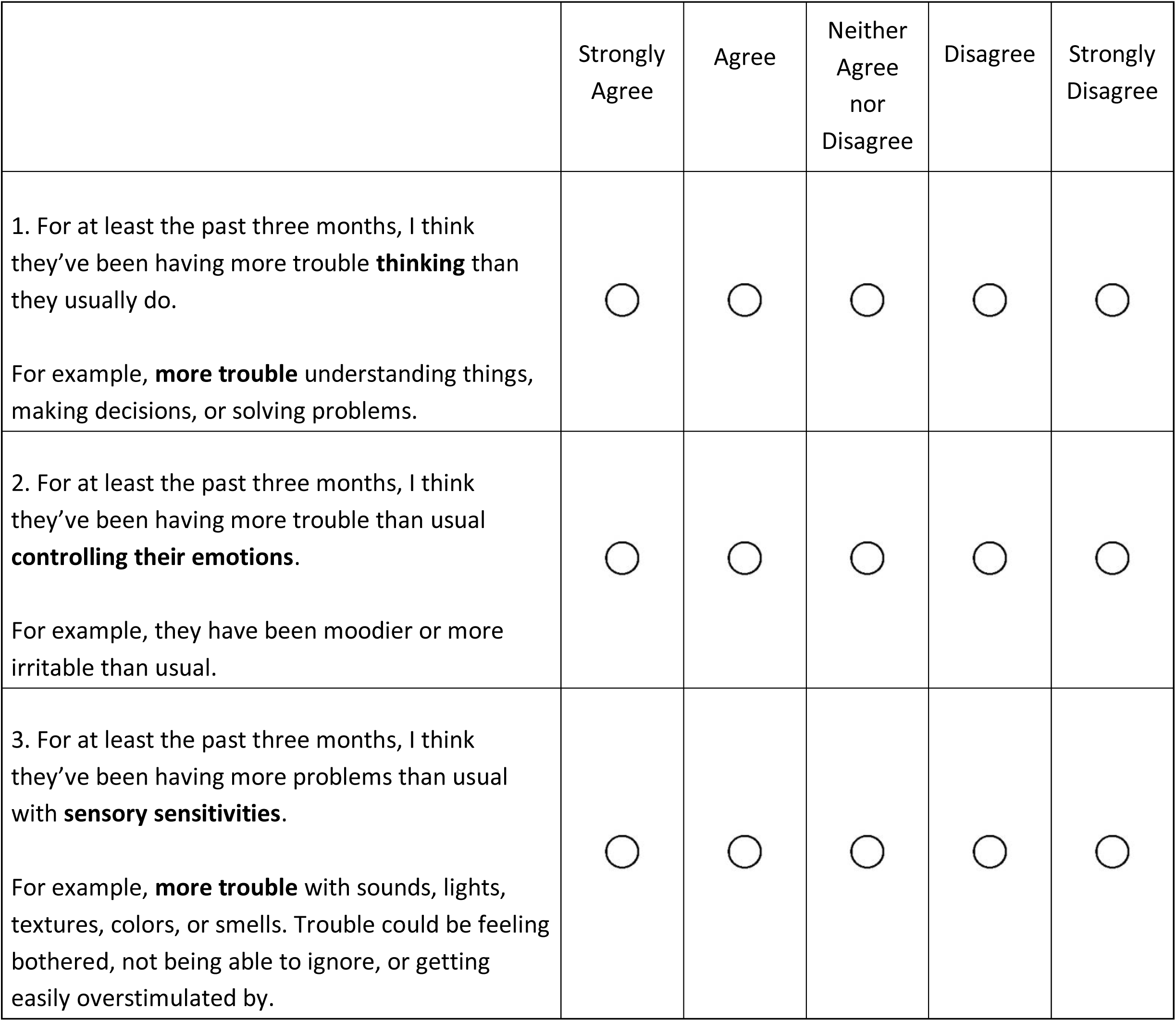
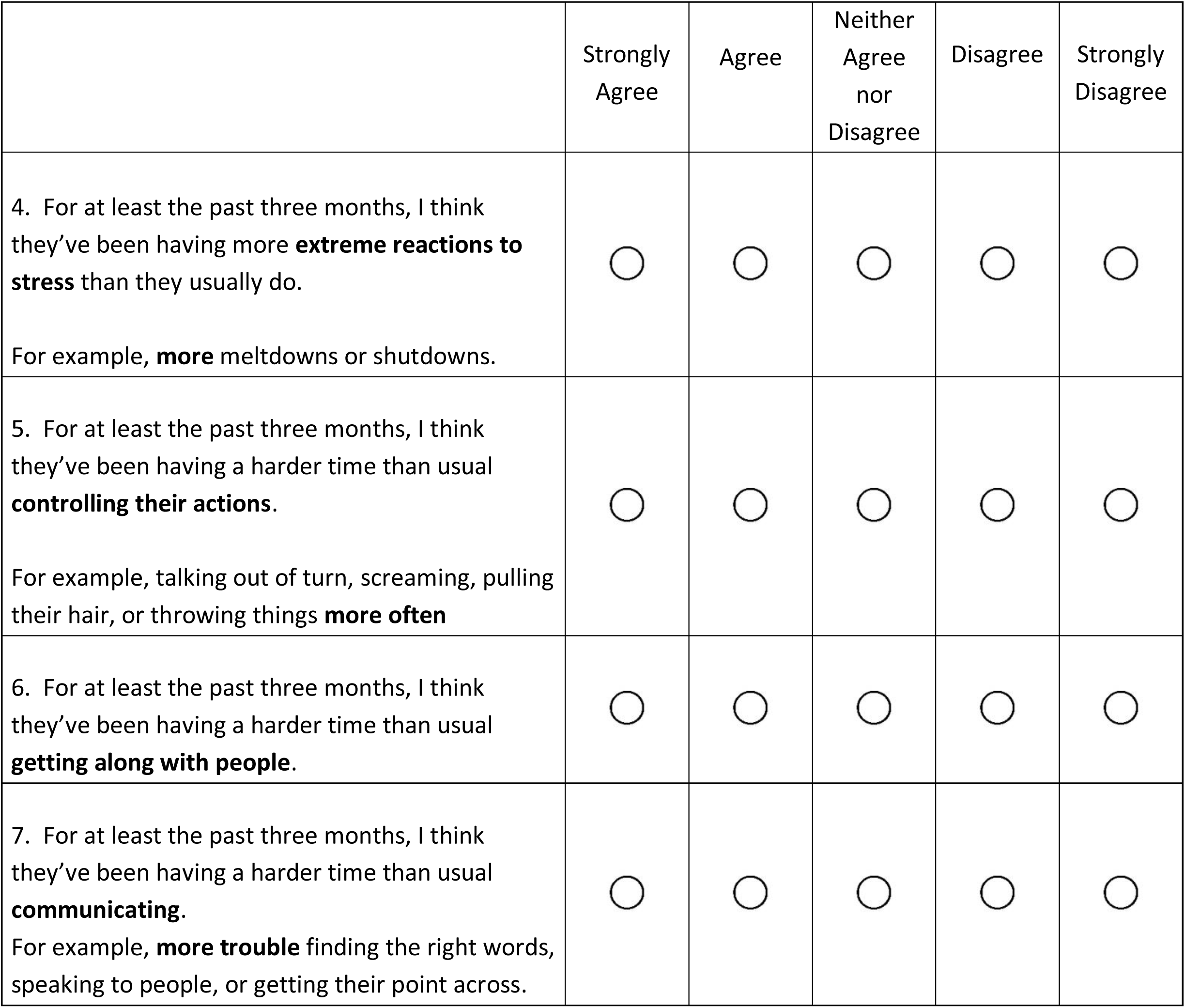
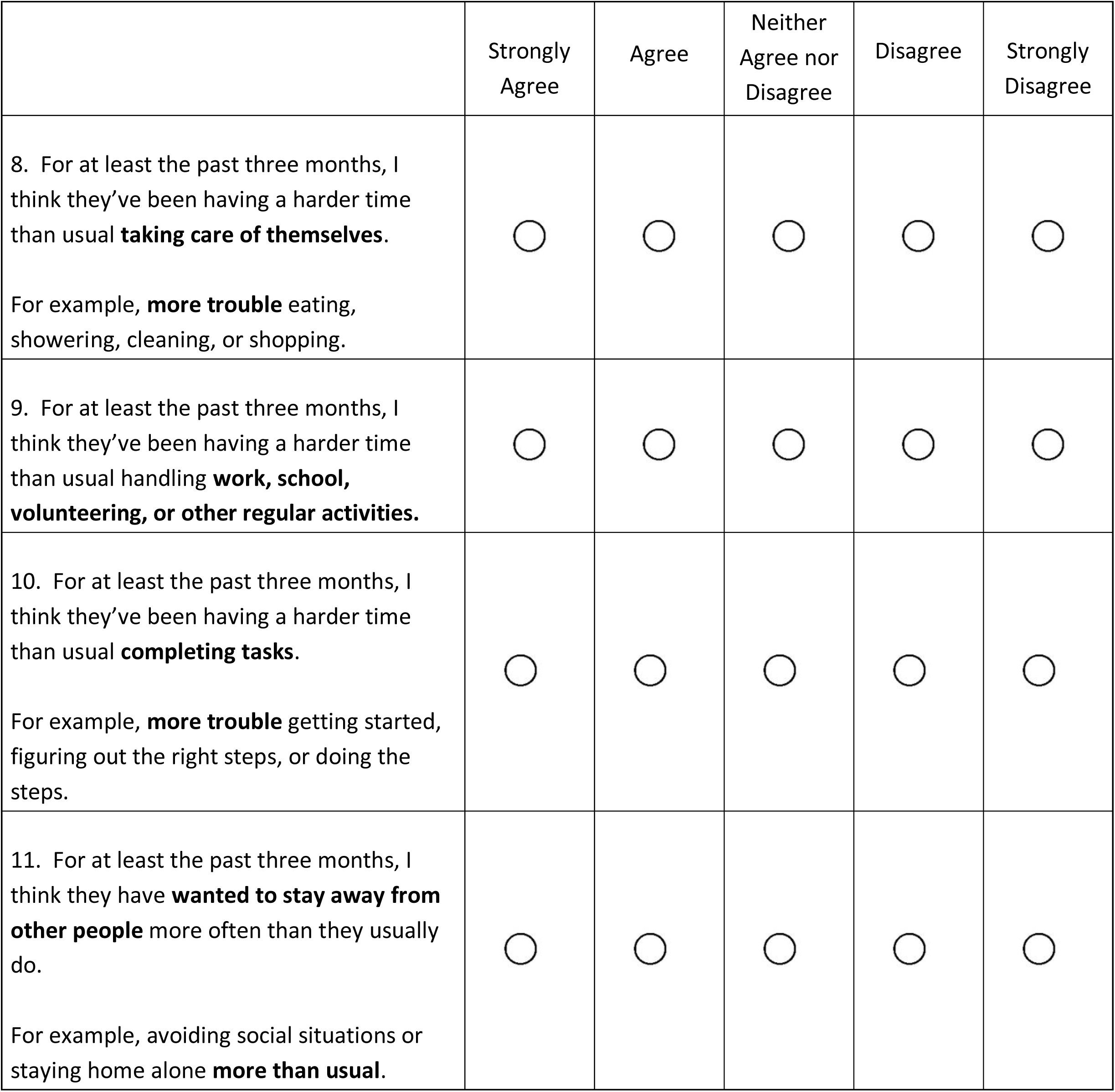
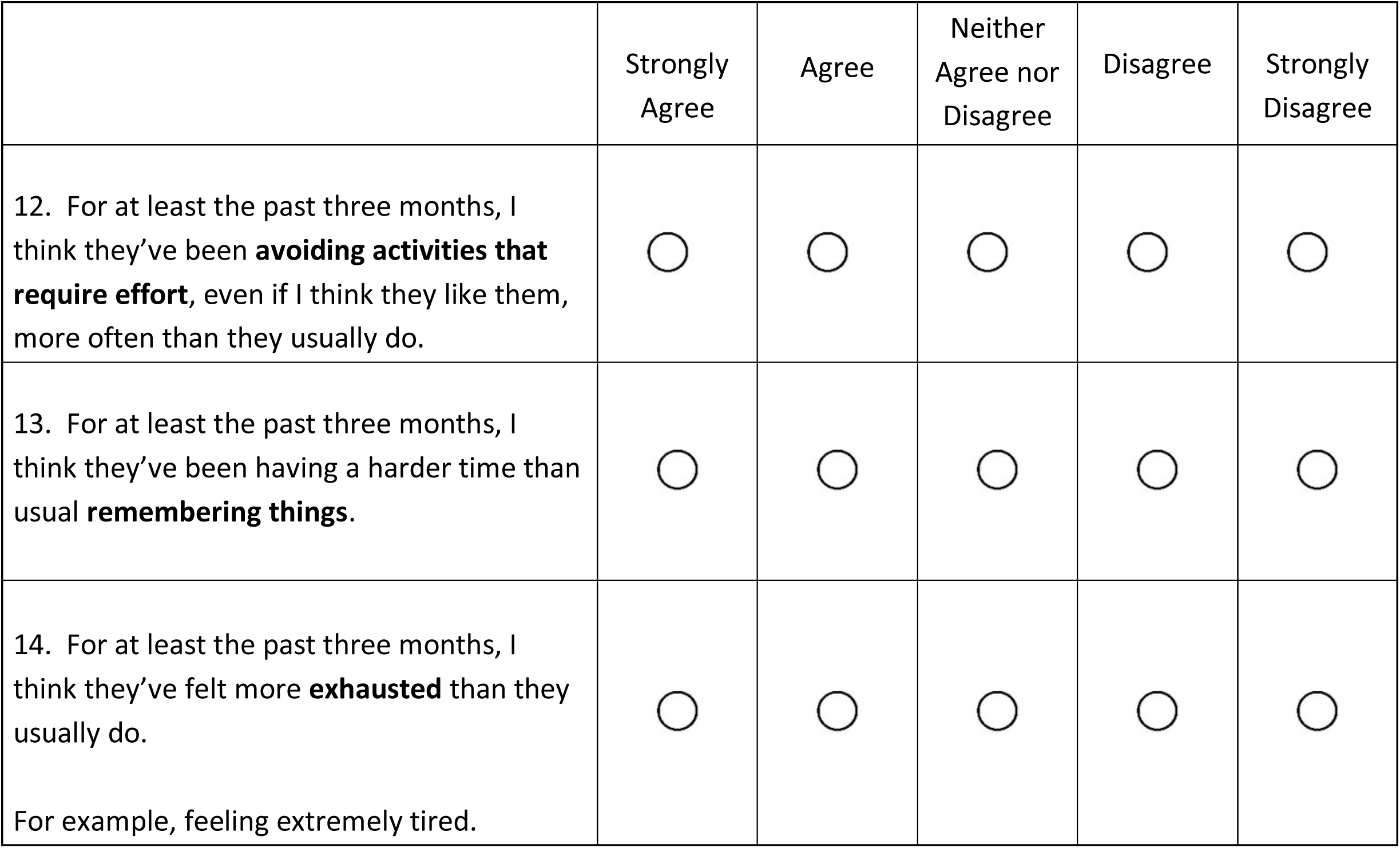

|  | Strongly Agree | Agree | Neither Agree nor Disagree | Disagree | Strongly Disagree |
| --- | --- | --- | --- | --- | --- |
| 1. For at least the past three months, I think they've been having more trouble **thinking** than they usually do.<br>For example, **more trouble** understanding things, making decisions, or solving problems. | ☐ | ☐ | ☐ | ☐ | ☐ |
| 2. For at least the past three months, I think they've been having more trouble than usual **controlling their emotions**.<br>For example, they have been moodier or more irritable than usual. | ☐ | ☐ | ☐ | ☐ | ☐ |
| 3. For at least the past three months, I think they've been having more problems than usual with **sensory sensitivities**.<br>For example, **more trouble** with sounds, lights, textures, colors, or smells. Trouble could be feeling bothered, not being able to ignore, or getting easily overstimulated by. | ☐ | ☐ | ☐ | ☐ | ☐ |
| 4. For at least the past three months, I think they've been having more **extreme reactions to stress** than they usually do.<br>For example, **more** meltdowns or shutdowns. | ☐ | ☐ | ☐ | ☐ | ☐ |
| 5. For at least the past three months, I think they've been having a harder time than usual **controlling their actions**.<br>For example, talking out of turn, screaming, pulling their hair, or throwing things **more often** | ☐ | ☐ | ☐ | ☐ | ☐ |
| 6. For at least the past three months, I think they've been having a harder time than usual **getting along with people**. | ☐ | ☐ | ☐ | ☐ | ☐ |
| 7. For at least the past three months, I think they've been having a harder time than usual **communicating**.<br>For example, **more trouble** finding the right words, speaking to people, or getting their point across. | ☐ | ☐ | ☐ | ☐ | ☐ |
| 8. For at least the past three months, I think they've been having a harder time than usual **taking care of themselves**.<br>For example, **more trouble** eating, showering, cleaning, or shopping. | ☐ | ☐ | ☐ | ☐ | ☐ |
| 9. For at least the past three months, I think they've been having a harder time than usual handling **work, school, volunteering, or other regular activities**. | ☐ | ☐ | ☐ | ☐ | ☐ |
| 10. For at least the past three months, I think they've been having a harder time than usual **completing tasks**.<br>For example, **more trouble** getting started, figuring out the right steps, or doing the steps. | ☐ | ☐ | ☐ | ☐ | ☐ |
| 11. For at least the past three months, I think they have **wanted to stay away from other people** more often than they usually do.<br>For example, avoiding social situations or staying home alone **more than usual**. | ☐ | ☐ | ☐ | ☐ | ☐ |
| 12. For at least the past three months, I think they've been **avoiding activities that require effort**, even if I think they like them, more often than they usually do. | ☐ | ☐ | ☐ | ☐ | ☐ |
| 13. For at least the past three months, I think they've been having a harder time than usual **remembering things**. | ☐ | ☐ | ☐ | ☐ | ☐ |
| 14. For at least the past three months, I think they've felt more **exhausted** than they usually do.<br>For example, feeling extremely tired. | ☐ | ☐ | ☐ | ☐ | ☐ |

### Additional Items

*The following items were also included in the Outcomes Project section on autistic burnout:*

#### Onset

*<If ‘Agree’ or ‘Strongly Agree’ options are selected for one or more items, Questions **#15** and **#16** are displayed>*

15. We are sorry to hear that they are experiencing some of the changes described above.

**How quickly** did these changes seem to come on?

◦ I think that things got worse for them within **days or weeks** (for example: days or a few weeks after an illness or major life event, whether the even was good or bad).
◦ I think that things **slowly built up over many months or years**.
◦ Other *<write-in>*

What do you think **may have brought on** the changes described in this section? (Check all that apply.

❑ A **physical illness, surgery, or accident** <u>(click to see examples)</u>
❑ A **major life event or transition** (whether it was a good or bad thing) <u>(click to see examples)</u>.
❑ A **long-term mismatch** between expectations and your abilities, resources, and supports <u>(click to see examples)</u>.
❑ Other *<write-in>*

#### Self-Reflection

Many autistic people have talked about something called “autistic burnout.”

17. Before taking this scale, had you heard of something called “autistic burnout”?

◦ Yes, and I feel like I have a good idea of what it is
◦ Yes, but I don’t know much about it
◦ No

18. Do you think they have **ever** experienced autistic burnout?

◦ Yes
◦ No
◦ I do not know

19. Do you think they are experiencing autistic burnout **now**?

◦ Yes
◦ No
◦ I do not know

#### Standard Caregiver Report Items on Perceived Change and Knowledge Sources

Overall, in the last six months, did the things this survey asked about…

◦ Get worse
◦ Stay about the same
◦ Get better

How do you know this information?

❑ They told me (in whatever mode they communicate).
❑ I have observed the action, behavior, or events.
❑ I can interpret their behaviors and/or vocalizations.
❑ Someone else told me.
❑ I feel like I can guess because I have known them for a long time.
❑ I feel like I can guess because I spend a lot of time with them.
❑ Other *<write-in>*
❑ None of the above

On a scale of 0 to 10, how confident are you in your responses?

*<0 - Not at all Confident to 10 - Totally Confident>*

OPTIONAL: If you are having trouble answering these questions, you may use this space to further clarify or explain your answers. *<write-in>*

### Hyperlinks

#### How to compare experiences to what is “usual” for them

- “Usual” means whatever you consider **normal for them**, or to be most **typical for them**. This may or may not be what’s true for them currently.
- If you think they have a lot of variation in these things, that is okay. Think about whether there’s a difference between the current variation and the usual variation.

#### Examples of how different people may answer these questions

- Loud sounds have always bothered Maria, but in the past, she could usually cope with it.

◦ Now it seems to be a lot worse.
◦ Her caregiver doesn’t remember exactly when Maria started having more trouble tolerating sounds, but she thinks it’s been gradually getting worse since she moved into a new group home about a year ago.
◦ She should answer “Strongly Agree” to the question about having more problems than usual with sensory sensitivities.
- Sam has always had a hard time controlling his emotions.

◦ It doesn’t seem like there has been any change in his ability to control his emotions.
◦ Sam’s caregiver should answer “Strongly Disagree” to the question about having more trouble than usual controlling emotions because the trouble hasn’t changed.
- Jamal usually has some trouble concentrating.

◦ Jamal has been sick the last 2 days. He is having a much harder time concentrating today than he usually does.
◦ Before getting sick, Jamal seemed to be concentrating about as well as he normally does.
◦ Jamal’s caregiver should answer “Strongly Disagree” to the question about having more trouble concentrating because the change hasn’t been going on for at least 3 months.

“A **physical illness, surgery, or accident** <u>(click to see examples)</u>.”

For example:

“A **major life event or transition** (whether it was a good or bad thing) (click to see examples).”

For example:

- Starting, graduating, or leaving a school, college, or training program
- Moving to a new home or living with different people
- Getting into a new romantic relationship, getting married, or starting a family
- Breaking up with someone or getting divorced
- Betting new caregivers or supporters
- A change in their job or employment
- Being in a really scary or dangerous situation
- The death of a loved one or the loss of a supporter

## Detailed Comparison Between Original and Adapted Version

Changes between the Original Autistic Burnout Scale and the Revised Autistic Burnout Scale-AASPIRE

|  | Original | Revised |
| --- | --- | --- |
| **Introduction** | The following questions ask you to compare what you are currently experiencing in your life with what you would consider to be your usual experiences.<br>“Currently” means within the past three months.<br>“Usual” means whatever you consider normal for you, or to be most typical for you.<br>This may be your current experience, or it may not be. If you have a lot of variation in your experiences with some of these things, that is okay. You can think about if there’s a difference between the current variation and the usual variation.<br>The questions ask you to pick an answer. Pick the best one you can; it does not need to be perfectly accurate. At the end of the questions, there is a write-in box where you can explain more if you felt any of your answers were not accurate. | The following set of questions focuses on **big, negative long-term** changes in your life.<br>• We are interested in things that you have been experiencing for **three months or longer**.<br>• Compare what you have been experiencing for three months or more to what is **usual for you**.<br>• If you have been feeling bad for a long time, think back to what things were like before you started feeling bad. |
| **Additional Information** | <No additional information in original> | • <u>What it means to have been experiencing something for three months or longer</u> |
|  |  | • <u>How to compare experiences to what is usual for you</u><br>• <u>Examples of how different people may answer these questions</u> |
| **Item Change #1** | 1. In the past three months, I've had more trouble thinking clearly than I usually do.<br>2. In the past three months, I've had a harder time making decisions for myself than I usually do<br>3. In the past three months, I've had a harder time deciding what is and is not important to pay attention to than I usually do.<br>4. In the past three months, I've had a harder time solving challenging problems than I usually do. | For at least the past three months, I've been having more trouble **thinking** than I usually do.<br>For example, **more trouble** understanding things, making decisions, or solving problems. |
| **Item Change #2** | In the past three months, I've been more moody than I usually am. | For at least the past three months, I've been having more trouble than usual **controlling my emotions**.<br>For example, I have been moodier or more irritable **than usual**. |
| **Item Change #3** | 1. In the past three months, I've had a harder time tolerating sensory input (for example, bright lights, loud sounds, or intense smells) than I usually do.<br>2. In the past three months, I've had a harder time preventing sensory overstimulation than I usually do.<br>3. In the past three months, I've had a harder time ignoring unimportant sensory input (for example, distracting sounds, sights, smells, or tactile sensations) than I usually do.<br>4. In the past three months, I've been avoiding stimulating environments, even if I like them, more often than I usually do. | For at least the past three months, I've been having more problems than usual with **sensory sensitivities**.<br>For example, **more trouble** with sounds, lights, textures, colors, or smells. Trouble could be feeling bothered, not being able to ignore, or getting easily overstimulated. |
| **Item Change #4** | 1. In the past three months, I've had more, or more severe, meltdowns than I usually do.<br>2. In the past three months, I've had more, or more severe, shutdowns than I usually do. | For at least the past three months, I've been having more **extreme reactions to stress** than I usually do.<br>For example, **more meltdowns** or **shutdowns**. |
| **Item Change #5** | In the past three months, I've had a harder time controlling my impulses than I usually do. | For at least the past three months, I've been having a harder time than usual **controlling my actions**.<br>For example, **talking out of turn, screaming, pulling my hair, or throwing things more often**. |
| **Item Change #6** | 1. In the past three months, I've had a harder time getting along with people I know well than I usually do.<br>2. In the past three months, I've had a harder time getting along with people at work, school, or in other community settings than I usually do. | For at least the past three months, I've been having a harder time than usual **getting along with people**. |
| **Item Change #7** | 1. In the past three months, I've had a harder time communicating my point to others than I usually do.<br>2. In the past three months, I've had a harder time finding the right words to communicate what I mean than I usually do. | For at least the past three months, I've been having a harder time than usual **communicating**.<br>For example, **more trouble** finding the right words, speaking to people, or getting my point across. |
| **Item Change #8** | In the past three months, I've had a harder time doing basic day-to-day activities<br>(for example eating, cleaning, shopping, or showering) than I usually do. | For at least the past three months, I've been having a harder time than usual **taking care of myself**.<br>For example, **more trouble** eating, showering, cleaning, or shopping. |
| **Item Change #9** | In the past three months, I've had a harder time managing work or school than I usually do. | For at least the past three months, I've been having a harder time than usual **handling work, school, volunteering, or other regular activities**. |
| **Item Change #10** | In the past three months, I've had a harder time managing the steps I need to take to complete tasks than I usually do | For at least the past three months, I've been having a harder time than usual **completing tasks**. |
|  |  | For example, **more trouble** getting started, figuring out the right steps, or doing the steps. |
| **Item Change #11** | 1. In the past three months, I've wanted to isolate myself from others more often than I usually do.<br>2. In the past three months, I've been avoiding social situations, even if I like them, more often than I usually do. | For at least the past three months, I have wanted to **stay away from other people** more often than I usually do.<br>For example, by avoiding social situations or staying home alone **more than usual**. |
| **Item Change #12** | In the past three months, been avoiding activities that require effort, even if I like them, more often than I usually do. | For at least the past three months, I've been **avoiding activities that require effort**, even if I like them, more often than I usually do. |
| **Item Change #13** | 1. In the past three months, I've had a harder time recalling things I know (long-term memory) than I usually do<br>2. In the past three months, I've had a harder time holding information in my mind for short periods of time (short-term or working memory) than I usually do. | For at least the past three months, I've been having a harder time than usual **remembering things**. |
| **Item Change #14** | 1. In the past three months, I've felt more mentally exhausted than I usually do.<br>2. In the past three months, I've felt more physically exhausted than I usually do. | For at least the past three months, I've felt more **exhausted** than I usually do.<br>For example, feeling **extremely tired**. |

**Supplement B: Participant Characteristics**

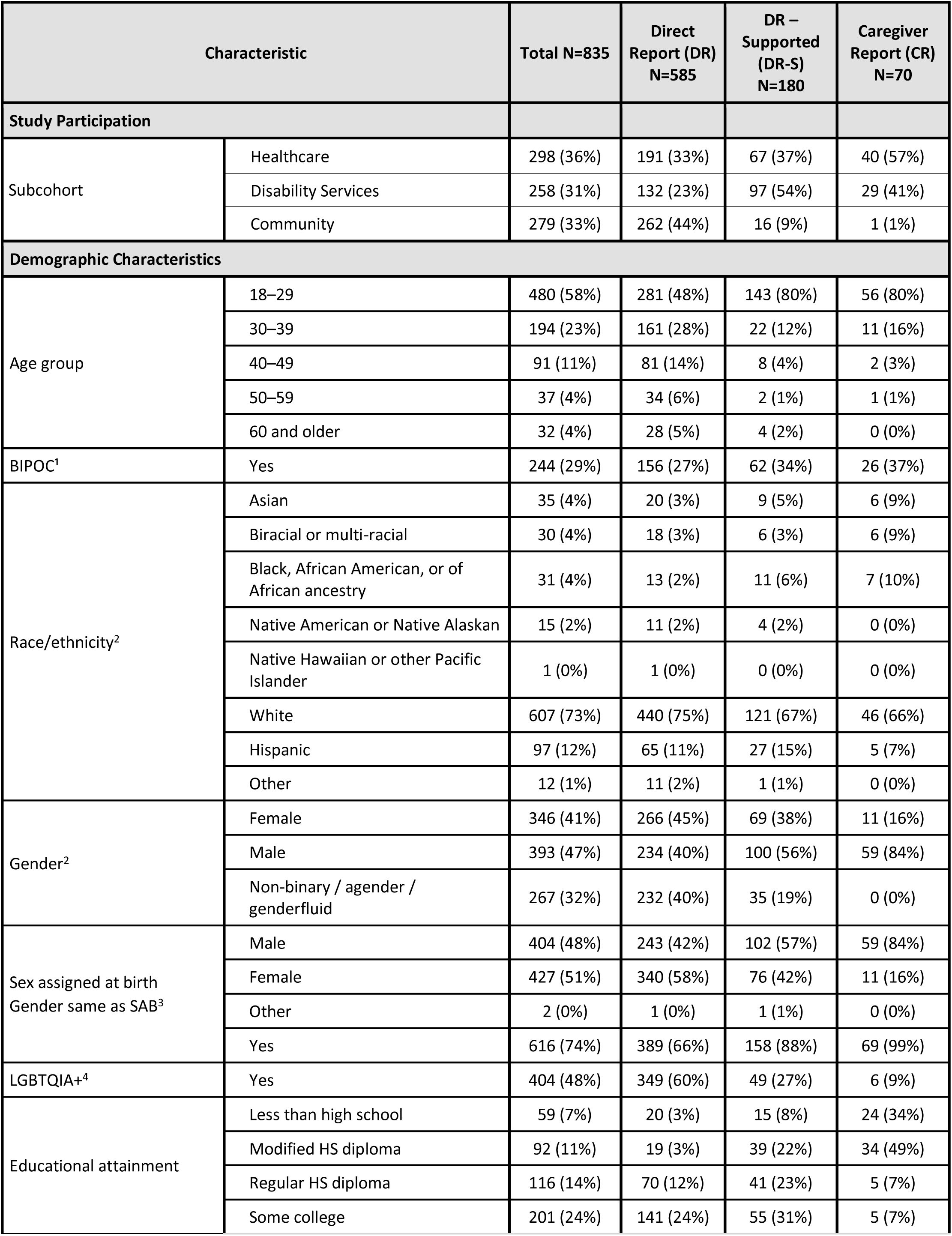
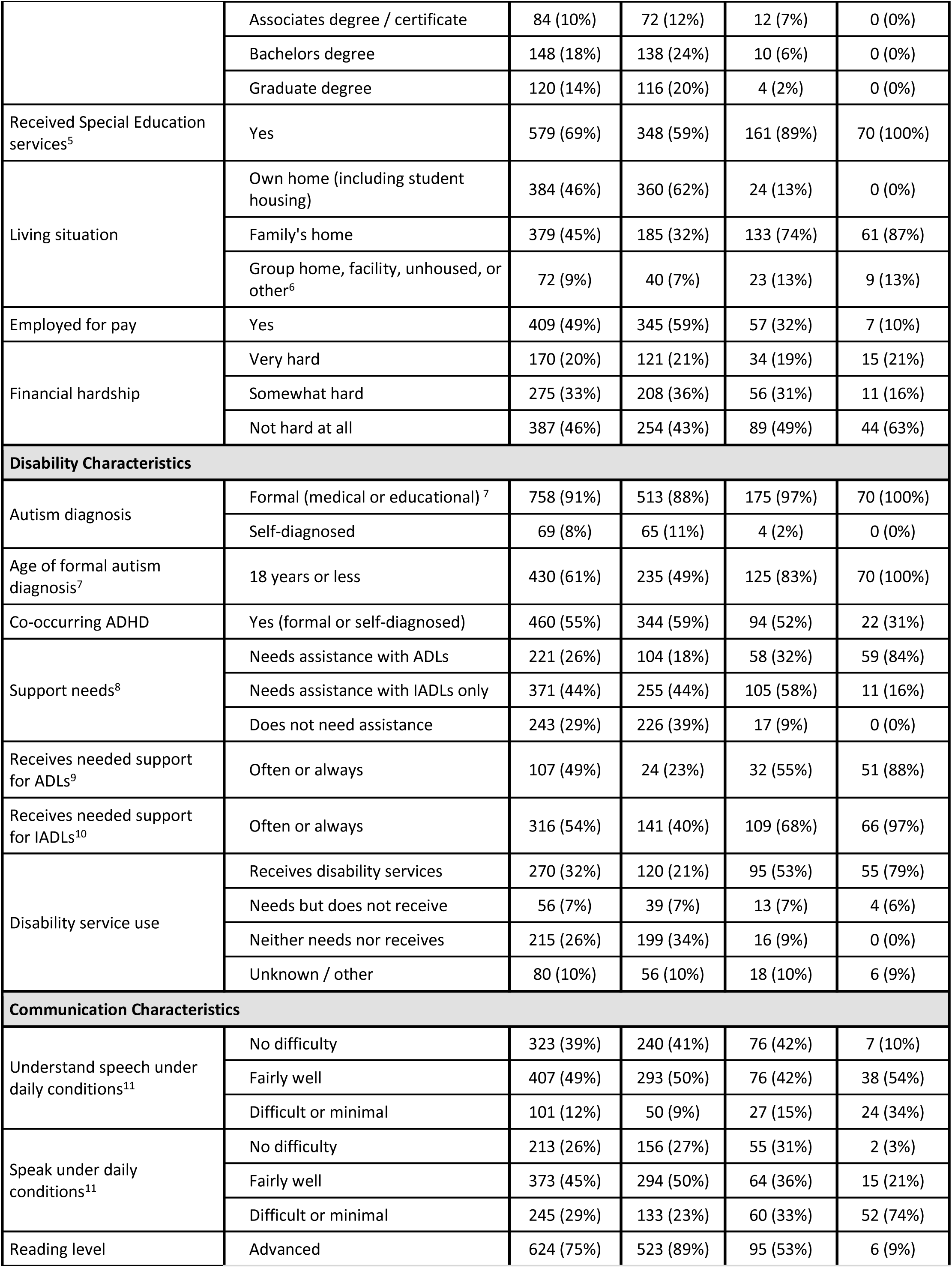
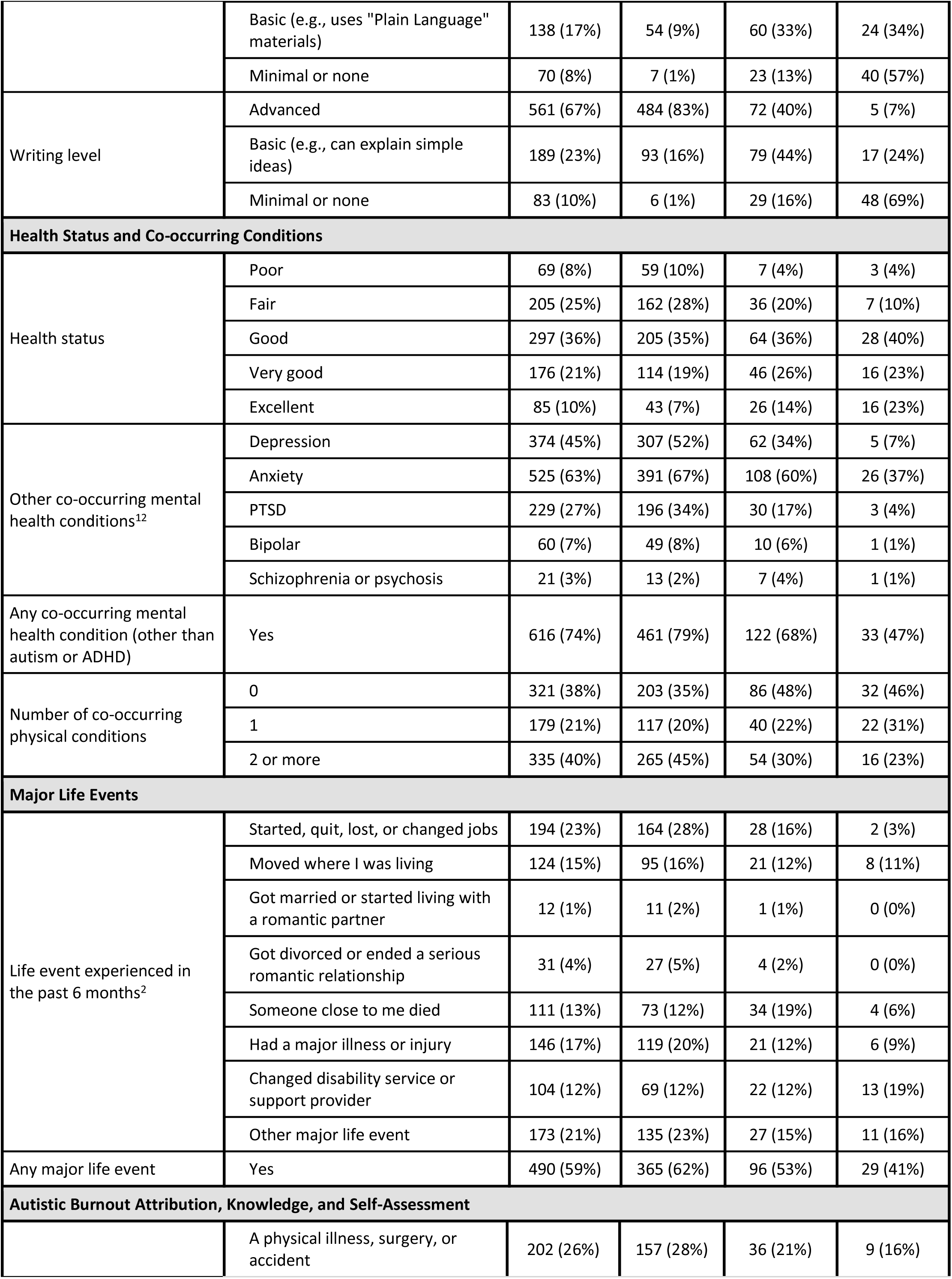
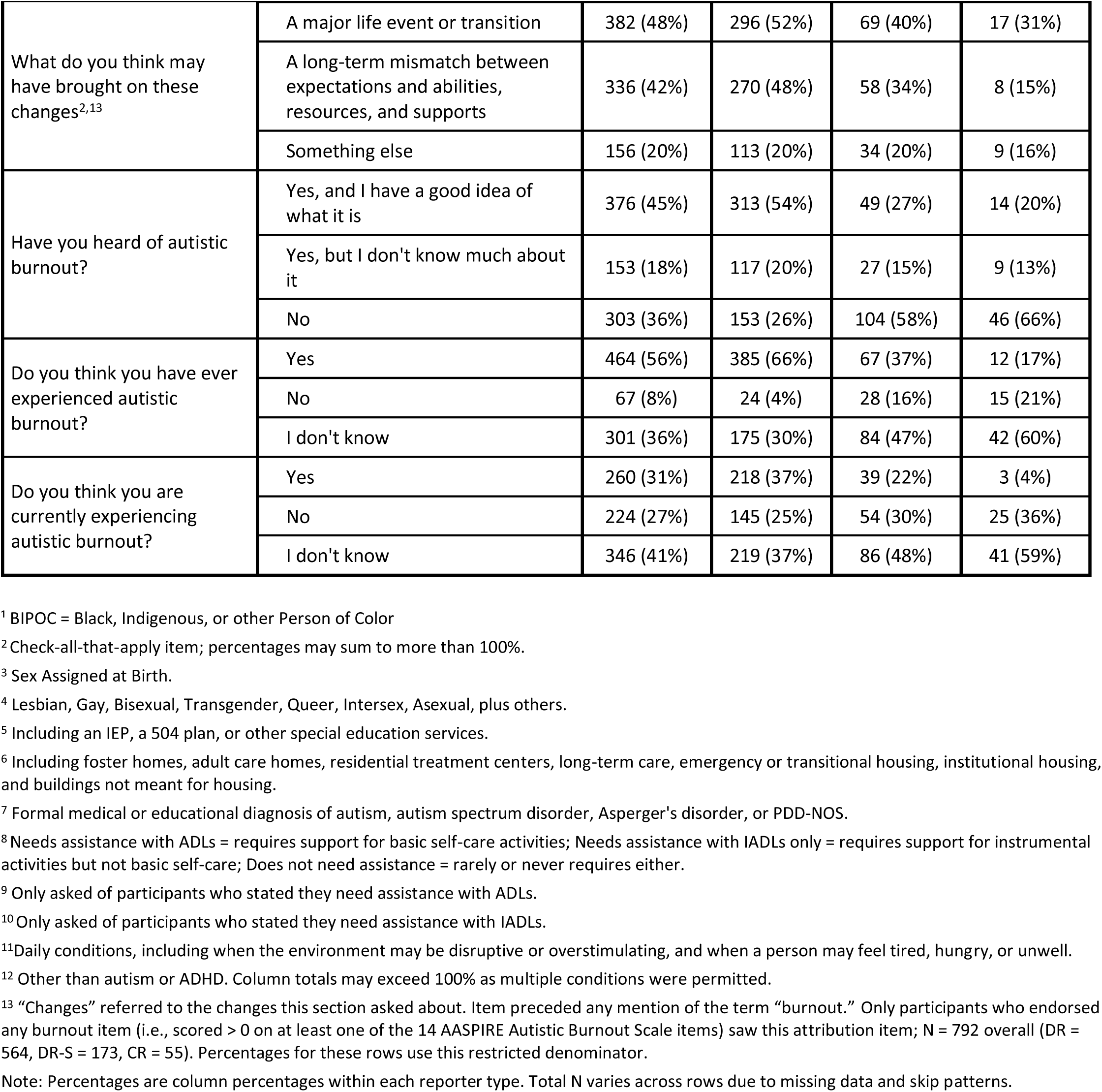

## Notes

### Competing Interest Statement

The authors have declared no competing interest.

### Clinical Protocols

https://doi.org/10.31234/osf.io/4m3bf_v6

